# iPlexD2Go Integrates AI-Guided Biomarker Panel Discovery with Decentralized Multiplex Diagnostics

**DOI:** 10.1101/2025.01.24.25321103

**Authors:** Zuan-Tao Lin, Chenling Tang, Yifei Kong, Yun Peng, Nianyu Jiang, Shuozhe Zhou, Yi Zhang, Sandeep Korupolu, Biswabandhu Jana, Shilin Gao, Yongli Li, Zhilong Wang, Cheng Liang, Quanwei Zhang, Youwen Liang, Ramesh Saxena, Wan Shou, Wojciech Matusik, Conor L. Evans, Tianfu Wu, Mei X. Wu

## Abstract

AI-guided biomarker panel discovery, multiplex detection, and automated assays have **each** advanced rapidly in the past decade, but their integration into cost-effective, user-friendly point-of-care (PoC) or home testing remains a critical translational barrier. Here, we develop a compact iPlexD2Go (intelligent mult<u>iplex</u> <u>d</u>iagnostic-<u>to-go</u>), an integrated platform enabled by the core multiplex sensing technology, **µChromaDot array (McDa)**. McDa could detect dozens of biomarkers simultaneously in a **one-biomarker-one-dot manner**, with **femtomolar-level analytical sensitivity** through a chemo-spatial amplification cascade, in which extensive chemical amplification on a 3D microneedle array was converted into 2D colorimetric readouts for seamless smartphone-based quantitative acquisition and analysis. Using clinical samples from patients with systemic lupus erythematosus (SLE; *n* = 42) and healthy controls (*n* = 34), we quantified five biomarkers and identified an optimized three-biomarker panel using a support vector machine (SVM) with 5-fold cross-validation. This panel discriminated SLE patients from healthy controls with 97% specificity and 93% sensitivity, representing a **70% improvement in specificity** over the clinical standard antinuclear antibody (ANA) test while maintaining the same sensitivity. We further engineered a mini-diagnostic device (miniDia) and companion App to automate McDa processing, smartphone-based image analysis, and user-friendly result reporting without laboratory infrastructure or trained personnel. By developing sensitive multiplex detection and integrating it with biomarker optimization and automated analysis, this end-to-end integrative iPlexD2Go provides an adaptable and cost-effective platform with strong potential for next-generation decentralized diagnosis, prognosis, and monitoring of various chronic diseases.

## Main

Point-of-care (PoC) testing becomes increasingly important for remote and decentralized healthcare, particularly with the steady growth of the aging population and the rapid expansion of digital healthcare^1–6^. Although PoC testing has been actively explored for decades, broadly accessible, multiplexed, user-friendly, and cost-effective PoC platforms remain critically lacking, making timely intervention difficult and increasing the risk of disease exacerbation. Disease pathogenesis is typically associated with intricate biological processes that encompass multiple signaling pathways. These processes often result in altered expressions of a panel of biomarkers, which can have either highly correlated, non-relevant, or mutually exclusive origins related to the underlying diseases. This occurs because our body strives to defend itself against abnormal attacks by various mechanisms, irrespective of their relevance. Thus, diagnosis, prognosis, treatment monitoring, and pathogenic processes could not be sufficiently determined by a single biomarker. A disease-specific biomarker panel is required for comprehensive insights into pathogenic processes^7,8^. However, both the routine approaches for multi-biomarker detection in clinics, including enzyme-linked immunoassay (ELISA), and other emerging technologies, such as microarrays, heavily depend on sophisticated and expensive instruments, for instance, plate readers and fluorescent scanners or microscopes, and well-trained technicians in fully equipped laboratories, especially for the low-abundance biomarkers. To translate multiplex detection capabilities from centralized laboratories into decentralized settings, enormous efforts have been devoted to developing PoC testing for multiplexed biomarker detection in the past two decades. Among them, colorimetric or fluorescent assays via imaging and analysis are increasingly attractive. Some recent assays have offered remarkable advances in sensitivity for detecting nucleic acids or small molecules^9,10^ or protein biomarkers^11–13^. With the development of miniature portable devices, microarray assays using fluorescent dyes have emerged as promising PoC platforms as well^13,14^. Yet, a portable and home-based **colorimetric** assay capable of multiplexed biomarker detection with high sensitivity and cost-effectiveness remains strongly desirable.

In the past two decades, biomarker panel discovery, multiplex sensing, and diagnostic assays have each advanced rapidly, but they have largely evolved as separate technological streams. Bridging these separate technological domains is essential for translating multiplex biomarker detection into practical point-of-care (PoC) or home testing. In this regard, advanced computational approaches, including machine learning (ML), have accelerated biomarker discovery and panel optimization, improving diagnostic performance across diverse clinical cohorts.^15–20^ However, these panels are typically selected based on statistical or biological performance without considering whether all selected biomarkers can be measured with sufficient sensitivity, dynamic range, and robustness in a practical decentralized setting. On the other hand, smartphone-enabled PoC platforms incorporating paper-based assays^21^, electrochemical sensors^22^, plasmonic microfluidic colorimetry assays^23^, wearable bioelectronic sweat sensors^24,25^, CRISPR-based wearable sensors^10^, or optical biosensors^26^ have enabled increasingly sensitive and multiplexed measurements, but generally detect predefined biomarkers rather than being designed to accommodate disease-specific biomarker panels. We therefore developed **iPlexD2Go** (intelligent multiplex diagnostic-to-go) to bridge the fundamental translational gap between identifying disease-specific biomarker panels and reliably and sensitively measuring them in practical, decentralized testing, as summarized in Fig. 1.

**Fig. 1.**
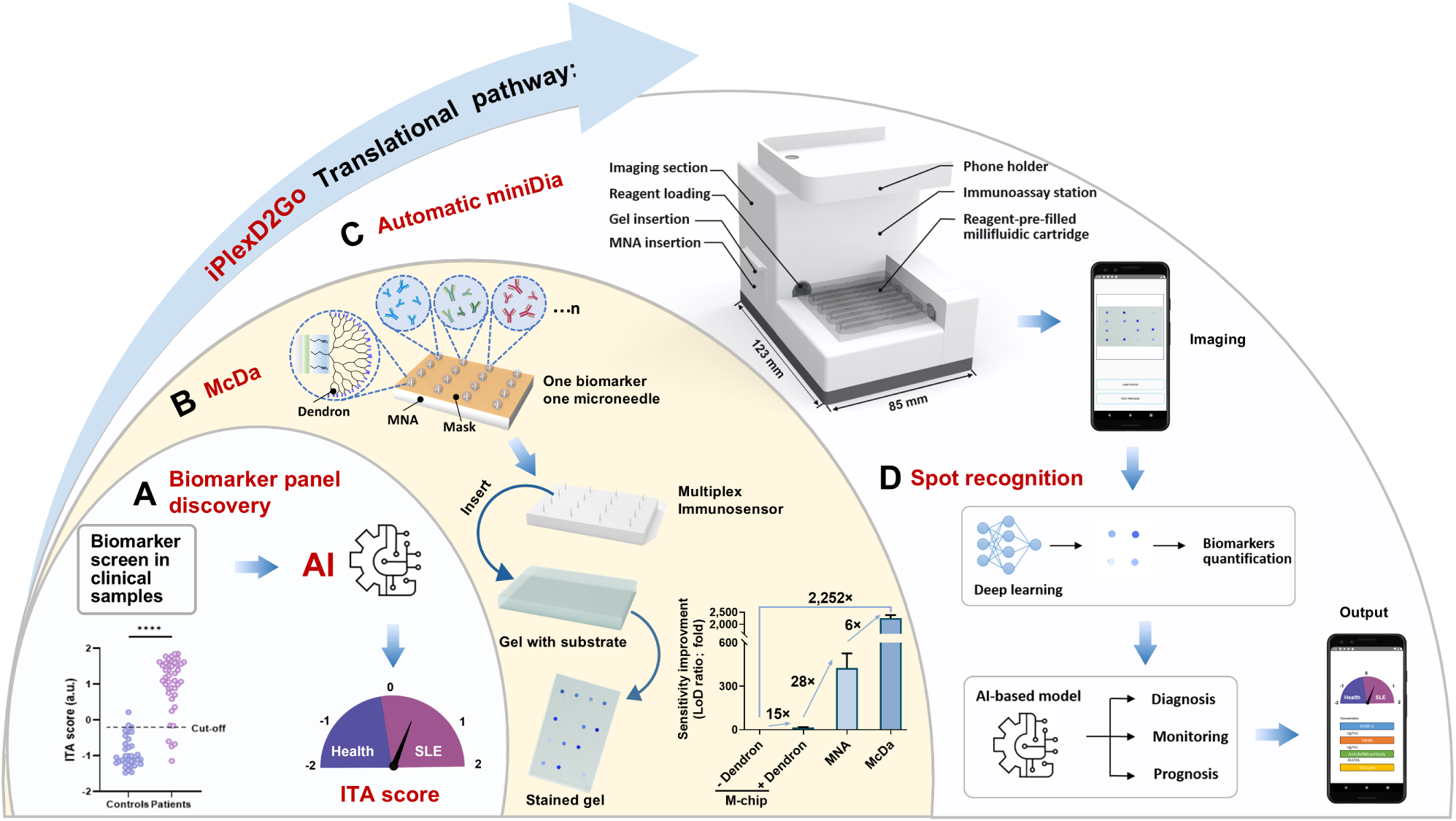
iPlexD2Go, intelligent multiplex diagnostic-to-go, closes the gap between AI-guided biomarker panel discovery and point-of-care testing (PoC). A. Clinical samples from healthy controls and patients with SLE were screened, identifying an optimal biomarker panel designed to an ITA score by machine learning modeling to significantly improve SLE diagnosis. B. The surface of a microneedle array (MNA) was chemically modified with PEI and dendrons to exponentially enrich amine groups for capture antibody conjugation. The MNA was covered by a mask (orange) perforated with a laser in a pattern precisely aligning with the microneedles in the MNA. These apertures each serve as a surface-tension-confined (STC) micro-reaction site to retain an isolated reaction droplet around a single microneedle and enable multiplex biomarker detection in a one-biomarker-one-microneedle fashion. After the immunoassay, an immunosensor with sandwich immunocomplexes formed on each microneedle was inserted into a substrate-saturated gel to convert the 3D signals into 2D signals for both signal intensification and cellphone-based data acquisition. These surface chemical modifications, 3D volumetric chromogenic precipitation around microneedles (spatial), and dimensional conversion (3D to 2D) together create a chem-spatial amplification cascade, resulting in a 2,252-fold improvement in sensitivity (Right, bottom). C-D. Schematic illustration of the portable device “miniDia” and data analysis. The device contains an immunoassay station, a reagent-pre-filled millifluidic cartridge, and an imaging station, together capable of executing the McDa assay automatically (C). An App was programmed to capture, recognize, and analyze individual spots in the stained gel to calculate the concentration of a specific biomarker by deep learning. An AI-based model was built with clinical sample analysis for disease diagnosis and monitoring at home or PoC, and for building a large database in the secure cloud center (D).

First, by leveraging ML algorithms, specifically a support vector machine (SVM) model with 5-fold cross-validation, we identified an optimized biomarker panel through screening of clinical samples. The AI-defined three-biomarker panel discriminated patients with systemic lupus erythematosus (SLE) from healthy controls with 97% specificity and 93% sensitivity (Fig. 1A), substantially outperforming the current clinical standard antinuclear antibody (ANA) test^27^. To enable the biomarker-panel analysis into adaptable, decentralized testing, we developed a micro-colorimetric assay, named a µChromaDot array (McDa), designed to measure a panel of biomarkers in a one-biomarker-per-dot format (Fig. 1B). The McDa strategically integrated surface-area amplification from a 3D microneedle array (MNA) with dendron decoration, resulting in a more than **2,252-fold** increase in sensitivity compared to conventional 2D microarray chips (Fig. 1B, right, bottom). The key to enabling one-capture-antibody-per-microneedle functionalization is the use of a thermoplastic polyurethane (TPU) laser-perforated mask to cover the MNA base, effectively blocking non-specific binding on the base and preventing cross-reactions among nearby microneedles. This architecture allows a single MNA to simultaneously quantify dozens of high-and low-abundant biomarkers in a single assay at a single sample dilution. After forming sandwich immunocomplexes on individual microneedles, the MNA is inserted into a gel saturated with colorimetric substrate, generating volumetric colorimetric precipitates corresponding to the amount of immunocomplex present on each microneedle (Fig. 1B, left, bottom). This conversion of 3D MNA signals into a 2D gel pattern effectively eliminates the need for costly analytical instruments and enables smartphone-based data acquisition and analysis, in marked contrast to the complex, instrument-reliant readout system for fluorescence-based assays. To further transform this assay for home tests, we engineered an automated “all-in-one” mini-diagnostic immunoassay device (miniDia) (Fig. 1C). The handheld miniDia comprises an immunoassay station, a reagent-prefilled millifluidic cartridge, and an imaging module for automated analysis. The individual biomarkers on the gel are quantified through tailored neural network algorithms, enabling the results to be directly interpreted and stored locally and securely on the patient’s smartphone or transmitted to secure cloud databases and healthcare providers (Fig. 1D). Collectively, the iPlexD2Go provides an adaptable and cost-effective unified multiplex diagnostic platform by end-to-end integration of AI-defined biomarker panels, a conceptually novel McDa signal amplification strategy, automated multiplex immunoassays, smartphone-based analysis, and patient-friendly result readout, holding strong potential for home-based diagnosis, prognosis, and monitoring of various chronic diseases.

## Results

### Development of multiplexed McDa with high sensitivity and reliability

To achieve laboratory-level analytical performance for multiplex protein biomarkers in a **laboratory**-**free** setting, we sought to develop an ultrasensitive, simple, cost-effective microassay, with sensitivity and reliability comparable to or exceeding those of widely used laboratory immunoassays in **clinical** practice, such as ELISA.^28–34^ We focused on **colorimetric**, rather than fluorescent, detection because of its low-cost, safer, and simpler readout without the excitation optics or laser-based instrumentation typically required for fluorescence detection. It is also readily compatible with smartphone-based signal acquisition and analysis. We first benchmarked the analytical sensitivity of several microarray-based assays against conventional ELISA using C-reactive protein (CRP) as a model biomarker. A protein microarray chip (flat dot; M-chip) was generated using a polymethyl methacrylate (PMMA)-chip with a planar surface immobilized with capture elements in the array. The PMMA-chip was prepared by casting a PMMA solution in a female polydimethylsiloxane (PDMS)-chip mold (Supplementary Fig. S1A). To enable multiplex detection in a single chip, we employed a laser to punch an array of microapertures, each with a diameter of 210 μm, on a mask of a 330 μm-thick TPU film with strong acrylic adhesion (Supplementary Fig. S1B-D). When the mask adhered to the surface of PMMA chip, each tiny aperture in the mask formed a surface-tension-confined (STC) droplet anchoring the aperture due to the hydrophobic and low-surface-energy properties of the mask. Each STC microdroplet functioned as a wetting barrier and held approximately 1 μL of a specific antibody/coupling reaction solution within each designated chip zone in the array, as illustrated in Fig. S1C and demonstrated with photos in Fig. S1D. This enabled individual capture elements to be mounted in spatially separated zones without cross-contamination or lateral spreading into neighboring zones. It also prevented the capture element solution from spreading across the basal area (Supplementary Fig. S1D). This simple yet ingenious design enabled multiplex detection by adding a single biomarker to each dot.

To immobilize a capture element on the chip, the surface of PMMA-chip was treated with oxygen plasma, coated with PEI, or both (Supplementary Fig. S1E). The same amount of CRP protein was immobilized on the three PMMA chips, followed by incubation with a detection antibody, streptavidin-HRP, and colorimetric substrate (Supplementary Fig. S1F). It was found that the one treated by both plasma and PEI had a higher signal intensity than the non-modified control or the chip treated by either alone (Supplementary Fig. S1G). To further enhance the immobilizing capture element, the surface of the PEI-coated M-chip was further decorated with different generations of dendrons (G2, G4, and G5) after the dendrimer was cut in half by tris(2-carboxyethyl)phosphine hydrochloride (TCEP) (Supplementary Fig. S1E, left). G5 dendrons exhibited the best performance, increasing the signal intensity by 225-fold in the presence vs. absence of a specific dendron, although G4 was also acceptable, with a 173-fold increase (Supplementary Fig. S1G). Hence, PMMA-chip modified with plasma, PEI, and G5 dendrons was used in subsequent studies.

CRP capture antibody at varying concentrations was immobilized on PMMA-chip (Fig. 2Ai), followed by a colorimetric microarray assay (Supplementary Fig. S1H). The chip with a clear background was obtained because of the mask-mediated blockade of non-specific binding (Fig. 2Aii). However, this protein microarray chip assay failed to achieve a desirable competitive limit of detection (LoD), about 7-fold less sensitivity than the gold-standard ELISA (Fig. 2Aiii and iv, 67.92 vs. 10.14 pg/mL). CRP is a highly abundant biomarker in serum (> 100 ng/mL), and its common clinical cut-off level is 40 µg/mL and can be readily obtained by the protein microarray chip^15^. Yet, this is not the case for low-abundant proteins (<100 ng/mL in serum), which are equally important as high-abundant biomarkers for many key disease biomarkers. To make things worse, at-home tests typically involve a small sample volume collected from a fingerstick, necessitating a higher dilution, such as 1:1,000, in contrast to the 10∼100-times lower sample dilution typically employed in laboratory tests. For instance, tumor necrosis factor receptor type II (TNFRII), which is expressed by T lymphocytes and can inhibit the activation of tumor necrosis factor α (TNFα), represents a low-abundant biomarker. Considering the clinical cut-off level of 1-3 ng/mL for TNFRII^35,36^, a 1:1,000 dilution for home tests reduced the concentration to 1-3 pg/mL, which is the minimal sensitivity required for clinical diagnosis. This level of sensitivity was barely reached by ELISA, with the LoD at 3.75 pg/mL (Fig. 2B), but it was far below the sensitivity achievable by the aforementioned microarray chip assay.

**Fig. 2.**
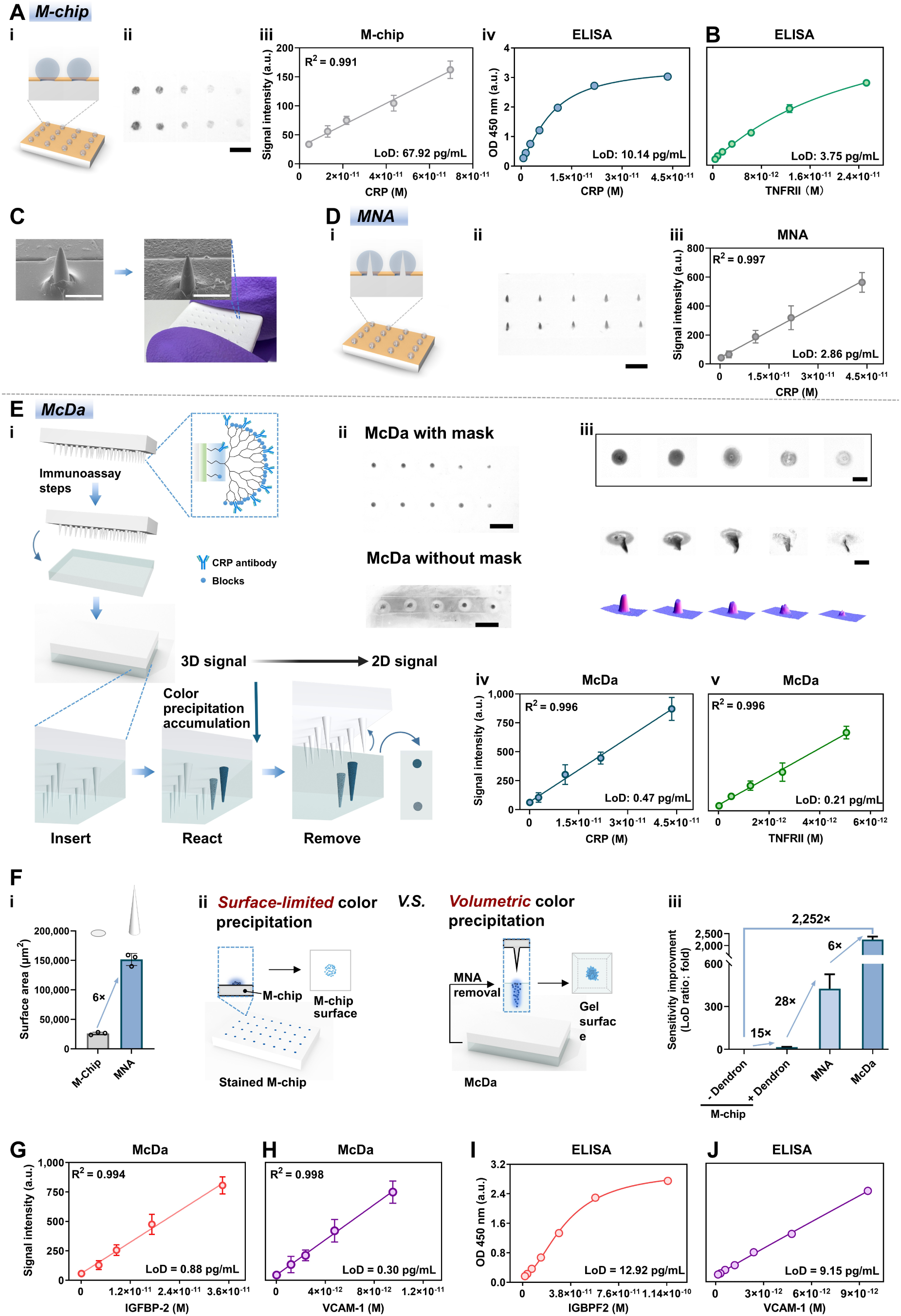
McDa development. **A-E**. A series of modifications results in drastic improvement in the specificity and sensitivity. **A**. **Microarray chip (M-chip) assay:** A laser-perforated mask (orange) was affixed to the surface-modified PMMA planar chip to create an array of STC micro-reaction sites (**i**). Image of the microarray chip in response to increasing concentrations (ii) and the calibration curve (**iii**) and corresponding standard curve of ELISA for CRP detection (**iv**). Scar bar: 500 µm. **B**. Standard curve of TNFRII ELISA kit. **C**. MNA increases the surface area for capture element immobilization relative to the M-chip in A. Photo of a MNA held by two fingers (bottom, right), and its SEM images before (left) and after (right) chemical surface modification. Scale bar, 400 µm. **D**. The illustration of **MNA-based assaying** (**i**). Image of stained microneedles with increasing concentrations of human CRP from 62.5 to 1,000 pg/mL (**ii**) and its calibration curve (**iii**). Scar bar: 1,000 µm. **E**. **McDa assay**: A conversion of 3D MNA signals into 2D signals with signal intensifying (**i**). The images of the stained gel in response to an increasing concentration of human CRP from 62.5 to 1,000 pg/mL in duplicate (quantitative analysis) (**ii, upper**). For comparison, the same assay in (**ii, upper**) was carried out without a mask (**ii, bottom**). Scale bar, 1,000 µm. Note, strong background staining was observed in the result obtained without a mask. One row of the representative gel is enlarged (**iii, upper**). Scale bar, 200 µm. The side-view image of the substrate precipitate distribution of the spots mirroring microneedles in the gel (**iii, middle**). Scale bar, 300 µm. The surface plot of the corresponding spot images in (**iii, upper**) using the Interactive 3-D Surface Plot plugin of ImageJ (**iii, bottom**). Calibration curve of McDa for CRP detection (i**v**) and TNFRII detection (**v**). **F**. Illustration of signal amplification cascade from M-chip, MNA, to McDa. Enlargement of the surface area of the MNA relative to spots (**i**). Comparison of surface-limited color precipitation on surface of M-chip and volumetric color precipitation in gel of McDa (**ii**) Dendron decoration provided a 14.85-fold enhancement in sensitivity through chemical amplification, whereas the 3D microneedle architecture and subsequent dimensional signal conversion in McDa provided an additional 146.65-fold (27.67 × 5.30) enhancement through spatial amplification and 3D to 2D signal conversion, respectively. (**iii**). McDa exhibits a lower LoD, compared to the microarray assay and directly staining MNA (**iii**). **G-H**. Calibration curves of McDa for IGFBP2 and VCAM-1 detection. **I-J**. Standard curves of commercial IGFBP2 and VCAM-1 ELISA kits. n= 3 independent tests each in triplicate. Data show mean ± s.d. a.u., arbitrary units.

To increase sensitivity, we strategically transitioned from a conventional 2D protein microarray chip to a PMMA-microneedle array (MNA), which substantially increased the surface area through a 3D architecture (Supplementary Fig. S2A). As shown in the scanning electron microscope (SEM) images (Supplementary Fig. S2B and Fig. 2C), the cone-shaped microneedle measured 500.00 ± 16.27 µm in length, with tip and base diameters of 10.20 ± 0.69 µm and 181.00 ± 6.78 µm, respectively. This 3D configuration increased the available surface area by 5.64-fold compared with the flat 2D microarray surface, with a potential for signal amplification by more than 1,000-fold (225 x 5.6) compared to a traditional flat 2D microarray chip (Supplementary Fig. S2C). The surface of PMMA-MNA was chemically modified, dendron-decorated, and masked as above (Supplementary Fig. S3A), followed by incubation with CRP capture antibody at varying concentrations in one-sample-per-microneedle fashion (Supplementary Fig. S3B and Fig. 2Di). Briefly, a TPU mask was perforated with an aperture array aligned to the MNA pattern and adhered to the MNA base (Fig. 2Di and Supplementary Fig. S3B). Approximately 1 µL of capture antibody reaction mixture was carefully pipetted onto each designated microneedle. The TPU surface confined the microdroplet locally and prevented spreading or coalescing with neighboring droplets owing to its intrinsic hydrophobicity and low surface energy (Fig. 2Di and Photos in Supplementary Fig. S3C, bottom). Each aperture thereby functioned as an independent STC micro-reaction site, enabling multiplexed antibody immobilization on a single MNA.

Successful coating of PEI and dendrons on the MNA surface was evidenced by the transition from a smooth PMMA-MNA surface to a coarse surface (Fig. 2C). After the formation of the HRP-sandwich immunocomplex on individual microneedles, the substrate solution was added directly onto the microneedle surface, and the resultant colorimetric intensity was directly measured microscopically on individual microneedles (Fig. 2Dii). Compared with the corresponding 2D-flat-dot array, the MNA platform achieved a 20-fold lower LoD (2.86 pg/ml in Fig. 2Diii vs 67.92 pg/ml in Fig. 2Aiii). However, direct imaging of stained microneedles required a sophisticated tunable stage to maintain each microneedle in sharp focus for accurate quantification by a microscope, representing a major barrier for PoC or home-based applications. In addition, owing to the spatial arrangement and 3D geometry of the individual microneedles, it was almost impossible to capture all microneedles simultaneously from the same perspective within a single image. Hence, data acquisition had to be performed one microneedle at a time, making the process very time-consuming and constraining its PoC potential.

To overcome these limitations, we innovatively introduced a gel matrix for signal transfer and amplification, named the µChromaDot assay (McDa). As depicted in Fig. 2Ei and Supplementary Fig. S4, the gel was pre-saturated with a substrate and subsequently penetrated by the MNA carrying HRP-sandwich immuno-complexes on individual microneedle surfaces. HRP catalysis generated blue precipitates around each microneedle within the gel (Supplementary Fig. S4, bottom). The precipitates accumulated vertically along the penetration depth of each microneedle, thereby amplifying the signal spatially within the gel. After removal of the MNA, the elastic gel rapidly resealed while preserving the precipitates in situ (Fig. 2Ei, right, bottom). Importantly, conversion of the 3D microneedle signals into 2D gel-based colorimetric spots dramatically enhanced signal intensity and enabled straightforward imaging from the gel surface (Fig. 2Ei, right, bottom, and Supplementary Fig. S5A). The ability of McDa to detect high and low abundant biomarkers was exemplified by representatives of CRP (> 100 ng/mL in serum), and TNFRII (< 100 ng/mL in serum). For CRP detection, duplicate CRP samples arranged in two MNA rows exhibited excellent reproducibility and specificity (Fig. 2Eii, upper). The gel background remained clear without nonspecific staining due to the TPU mask-assisted localization strategy. In contrast, direct deposition of reaction solution onto unprotected MNA surfaces caused droplet spreading and cross-contamination between adjacent microneedles, resulting in substantial nonspecific background (Fig. 2Eii, bottom). The resulting colorimetric spots were circular, approximately 207 µm in diameter, and their intensities correlated with CRP concentrations (Fig. 2Eiii, upper, and 2Eiv). Their size closely matched the microneedle base diameter, indicating minimal diffusion of precipitates within the gel. Notably, the precipitates formed cone-shaped structures mirroring the geometry of the microneedles, further contributing to signal enhancement (Fig. 2Eiii, middle and bottom). The G5 dendron decoration on PEI coated MNA surface achieved a further ∼28-fold increase in signal intensity compared to the surface without dendron (Supplementary Fig. S5B). Consequently, the McDa achieved an LoD of 0.47 pg/mL for CRP, representing a 147-fold increase in sensitivity compared with the conventional protein microarray chip and a 22-fold improvement over ELISA (Fig. 2Aiii vs. Fig. 2Eiv). The substrate-saturated gel was stable, and no significant change in signal was observed after it was sealed and stored at 4°C for 1, 3, and 6 months (Supplementary Fig. S6).

The dramatic gain in sensitivity likely stemmed from cumulative chem-spatial signal amplifications, integrating increased 3D surface area, multilayer surface modifications, and dimensional amplification through 3D-to-2D signal conversion, together yielding a 2,252-fold enhancement (Fig. 2Fiii). In addition to its ultrahigh sensitivity, McDa demonstrated a broad linear detection range of 0.98–1,000 pg/mL for CRP, exceeding that of ELISA (Fig. 2Eiv). Similar dose-dependent responses and highly linear standard curves were also observed for TNFRII detection (Supplementary Fig. S7 and Fig. 2Ev), with a LoD 17.86-fold lower than ELISA, confirming the capability of McDa to detect low-abundance biomarkers at clinically relevant levels. The substantial sensitivity enhancement enabled simultaneous measurement of multiple biomarkers from only 1 µL of diluted serum, readily obtainable by fingerstick sampling. For example, TNFRII in patient serum diluted 1:1000 was successfully quantified by McDa at concentrations of 5.45 ng/mL and 3.14 ng/mL (Supplementary Fig. S8A,B), whereas both conventional microarray assays and ELISA failed to detect TNFRII under the same dilution conditions.

### McDa measures disease biomarkers with higher sensitivity and broader linear ranges than ELISA

To demonstrate multiplexed quantifications of biomarkers in disease diagnosis, we selected five biomarkers according to biological pathways implicated in systemic lupus erythematosus (SLE) pathogenesis, including CRP, TNFRII, insulin-like growth factor binding protein-2 (IGFBP-2), vascular cell adhesion molecule-1 (VCAM-1), and anti-double-stranded DNA antibody (anti-dsDNA antibody). First, pairs of capture and detection antibodies for each biomarker were evaluated and compared between the McDa and commercially available ELISA kit side-by-side. Among the five biomarkers, the calibration curves of CRP and TNFRII are shown in Fig. 2Eiv&v. The calibration curves of human IGFBP-2 and VCAM-1 had the LoD of 0.8778 pg/mL (2.5214× 10^-14^ M), and 0.3001pg/mL (4.049 × 10^-15^ M), with the linear ranges from 2.34 to 1,200 pg/mL for IGFBP-2, and 0.98 to 1,000 pg/mL for VCAM-1, respectively, (Fig. 2G-H). The detection sensitivity for IGFBP-2 and VCAM-1 increased by 14.78-fold, and 30.48-fold, respectively, by McDa over commercial ELISA kits (Fig. 2I-J). The comparisons of commercial ELISA assays of human CRP, IGFBP-2, TNFRII, and VCAM-1 with McDa results were summarized in Supplementary Table S1. The LoD values of McDa colorimetric assays are at least one order of magnitude lower than those of ELISA kits, representative of 21-fold higher sensitivity on average than ELISA.

### Spot recognition by deep learning

The size and morphology of spots sometimes display irregularity in the gel owing to extremely high or low amounts of biomarkers bound to the microneedle and slight variations in microneedle insertion in McDa. For precise data acquisition, we adopted deep learning for boundary detection using U-Net architecture^37^. The network architecture consisted of two parts; a “contracting path” and an “expansive path”. Original spot images in the gel were shown in Supplementary Fig. S9 left, while the images processed by deep learning were shown in Fig. S9 right. The latter displayed spots with clear-defined boundaries, ensuring reproductivity and accuracy of data acquisition in McDa.

### Clinical evaluation of McDa for SLE diagnosis

Next, serum samples collected from patients with SLE (N = 42) and healthy controls (N = 34) were measured by the McDa to validate its clinical potential with ELISA running in parallel (Fig. 3A). A High correlation drew similar conclusion between the two assays (Fig. 3B): i.e., in comparison with healthy controls, levels of IGFBP-2, TNFRII, VCAM-1, and anti-dsDNA antibody rose significantly in SLE’s serum and especially, the levels of IGFBP-2, TNFRII, and anti-dsDNA antibody were significantly higher in patients with SLE than those in healthy controls. However, there was no significant difference in the level of CRP between lupus patients and healthy controls. We then performed a paired correlation analysis of the results between ELISA and McDa obtained from the same patient. As shown in Fig. 3C, strong correlations were found between the two assays, with R^2^ values of 0.9467, 0.9472, 0.9339, 0.9050, and 0.9467 for CRP, IGFBP-2, TNF-RII, VCAM-1, and anti-dsDNA antibody, respectively, suggesting the high reliability, sensitivity, and accuracy of the McDa for single biomarker detection comparable to clinical ELISA kits, yet, with 100x less serum samples and all reagents.

**Fig. 3.**
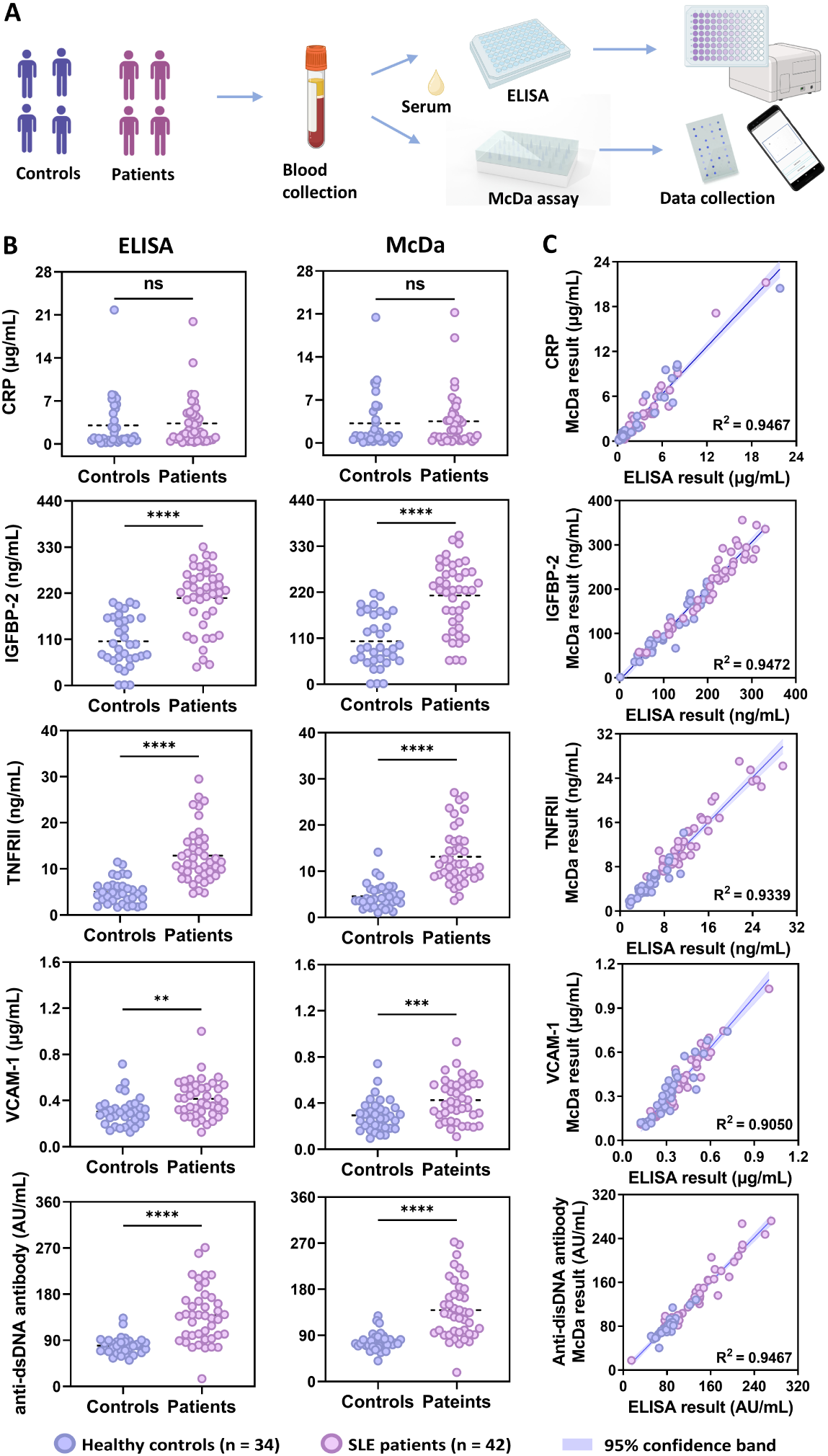
Clinical evaluation of selected biomarkers for SLE. **A.** Schematic illustration of measuring SLE biomarkers with McDa compared to ELISA side-by-side: created with BioRender.com. **B**. CRP, IGFBP2, TNFRII, and VCAM-1 and anti-dsDNA antibody were measured in serum samples from lupus patients (n = 42) and healthy controls (n = 34) by McDa and corresponding ELISA kits. **C**. The paired correlation test was carried out for the McDa versus ELISA results. Data show mean ± s.d. NS, not significant. Each symbol represents a single serum sample. ** P = 0.0016, *** = 0.0008, **** P< 0.0001 by two-tailed unpaired Student’s *t*-test. a.u., arbitrary units.

It is noteworthy that none of the five biomarkers alone could discriminate patients from healthy controls accurately and specifically, owing to considerable overlaps between SLE patients and healthy controls. Therefore, machine learning algorithms were employed to identify **a minimal number** of biomarkers for SLE diagnosis. To accomplish this, an immunosensor capable of measuring the five biomarkers simultaneously in a single sample at a single serum dilution had to be made, which turned out to be very challenging. Firstly, cross-reactivity is one of the major issues in multiplex biomarker detection. To address this challenge, we prepared a series of solutions containing one targeted biomarker at a fixed concentration while the other three biomarkers with increasing concentrations, followed by the assay. For all four biomarkers, when the targeted biomarker was fixed at 20 pg/mL, the signal intensity remained stable, and no significant signal intensity changes were observed with the increasing concentrations of the other three co-existed biomarkers, demonstrating negligible cross-reactivity of McDa for multiplex biomarkers detection (Fig. 4B). Next, thanks to the broad linear ranges of low abundant biomarkers IGFBP-2, VCAM-1, and TNFRII, all the samples could be measured by a single serum dilution of 1:1000. To accommodate the low and high abundant biomarkers of TNFRII and CRP with the same serum dilution, we reduced the amount of CRP capture antibody immobilized on the designated microneedles based on a needed sensitivity (Supplementary Fig. S10).

**Fig. 4.**
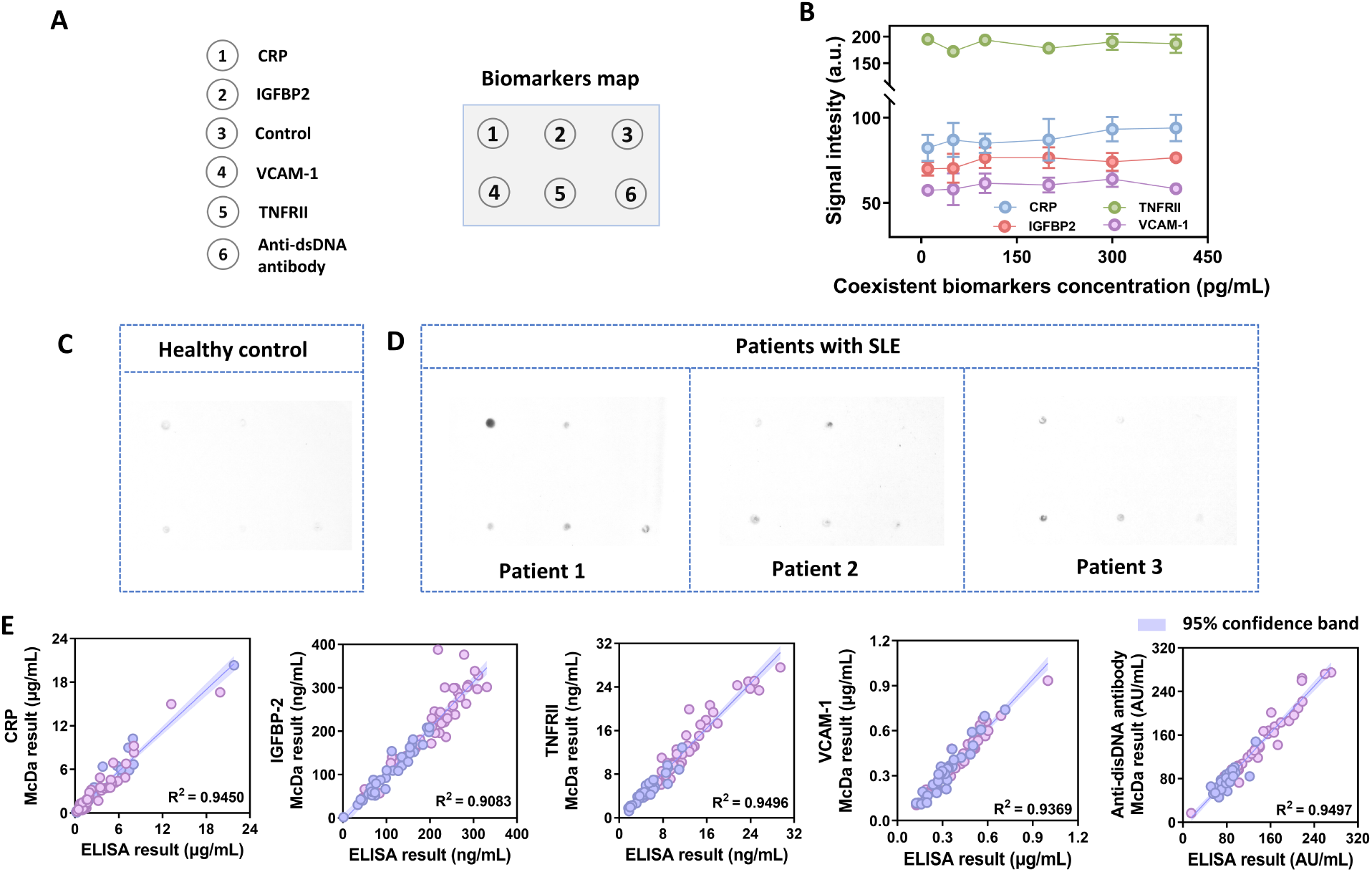
Validation of McDa assay for its clinical potential in multiplexed detection of biomarkers. **A.** An immunosensor with five indicated biomarkers and a control in designated locations of the array. For anti-dsDNA antibody measurement, the microneedle was conjugated with mBSA and then incubated with dsDNA. **B**. Plot suggests little cross-reactivity over a broad range of concentration. We fixed the concentration of the detected biomarker at 20 pg/mL while increasing the concentration of other biomarkers from 10 to 400 pg/mL for these tests. **C-D**. Representative gel images of healthy controls and SLE patients using a single immunosensor per patient or control. **E**. The paired correlation tests were conducted for McDa versus ELISA results. a.u., arbitrary units.

After the immobilization of different capture antibodies on designated microneedles in a single immunosensor, we went on to measure the five biomarkers on the serum samples collected from patients with SLE (N = 42) and healthy controls (N = 34) after 1:1,000 dilution. This was possible because the concentrations of all five biomarkers fell into the linear range of the standard curve of CRP, TNFRII, IGFBP-2, and VCAM-1, as shown in Fig. 2Eiv, v, G, and H. The representative McDa gel dot images of healthy controls (C) and 3 SLE patients (D) were shown in Fig. 4, which measured five biomarkers simultaneously in a single McDa gel for each patient sample. The corresponding concentrations of each biomarker in these control and patient serum samples were calculated from specifically stained dots in the gel by ImageJ and compared side-by-side with those attained by standard ELISA kits (Supplementary Table S2). Once again, a clear background was consistently observed in all samples, reaffirming the high specificity of McDa with negligible influence of other molecules in serum. Similar to the result of McDa for single biomarker detection, multiplex biomarker detection revealed strong correlations between the two assays, with R^2^ values of 0.9450, 0.9083, 0.9496, 0.9369, and 0.9497 for CRP, IGFBP-2, TNFRII, VCAM-1, and anti-dsDNA antibody, respectively, (Fig. 4E). Consistently, McDa outperformed the gold-standard ELISA in terms of reliability and accuracy, with higher sensitivity, cost-effectiveness, and readily result interpretation. We also compared the McDa results from multiplex biomarker detection with single biomarker detection, and found a strong correlation between them, with R^2^ values of 0.9506, 0.9238, 0.9403, 0.9386, and 0.9640, for CRP, IGFBP-2, TNFRII, VCAM-1, and anti-dsDNA antibody, respectively (Supplementary Fig. S11). The strong correlation demonstrates that the McDa offers excellent measurement stability between multi-biomarker detection and corresponding single-biomarker detection.

### Identification of a new SLE diagnosis score (ITA) by machine learning

To define an optimized biomarker panel for SLE diagnosis, we developed an AI model using support vector machines (SVM) algorithm to evaluate each biomarker either singly or in various combinations of the five biomarkers. A five-fold cross-validation method was also implemented to prevent overfitting and ensure generalizability (Fig. 5A). Table 1 summarizes the SLE diagnostic performance metrics of biomarkers detected by ELISA and McDa, including area under the curve (AUC), accuracy, sensitivity, and specificity. These metrics were reported as the average of all five fields during cross-validation for both training and testing sets. A final AI model was established using all the data to generate receiver operating characteristic (ROC) curves and calculate the AUC. The optimal cutoff values were obtained from ROC curves. The results showed that in both assays, the AUC values of IGFBP-2, TNFRII, and anti-dsDNA antibody were greater than 0.8, while the AUC values of CRP and VCAM-1 were lower than 0.7 (Table 1), in agreement with the finding in Fig. 3B. Remarkably, a biomarker panel with a combination of **I**GFBP-2, **T**NFRII, and **A**nti-dsDNA antibody exhibited the best performance. This combination, termed “ITA”, could serve as a new SLE diagnostic score. Compared with the single biomarker of IGFBP-2, TNFRII, or Anti-dsDNA antibody in ITA, ITA had a higher AUC value (Fig. 5 C&D). Among the test sets of the five individual biomarkers and their different combinations, ITA showed the highest AUC value with a minimal number of biomarkers in both ELISA and McDa assays (Table 1). ITA score with a cut-off value was -0.1753 between SLE patients and healthy controls with a significant difference, and only a few cases overlayed, demonstrating that the ITA score could distinguish SLE patients from healthy controls effectively (Fig. 5 E&F). The performance of McDa for multi-biomarker detection was comparable to that of ELISA kits (Fig. 5 E&F). The results demonstrate convincingly that more biomarkers do not necessarily lead to better diagnosis. The AI model proves to be very useful in determining the best biomarker set for disease diagnosis, a task that was previously impossible without AI model training.

**Fig. 5.**
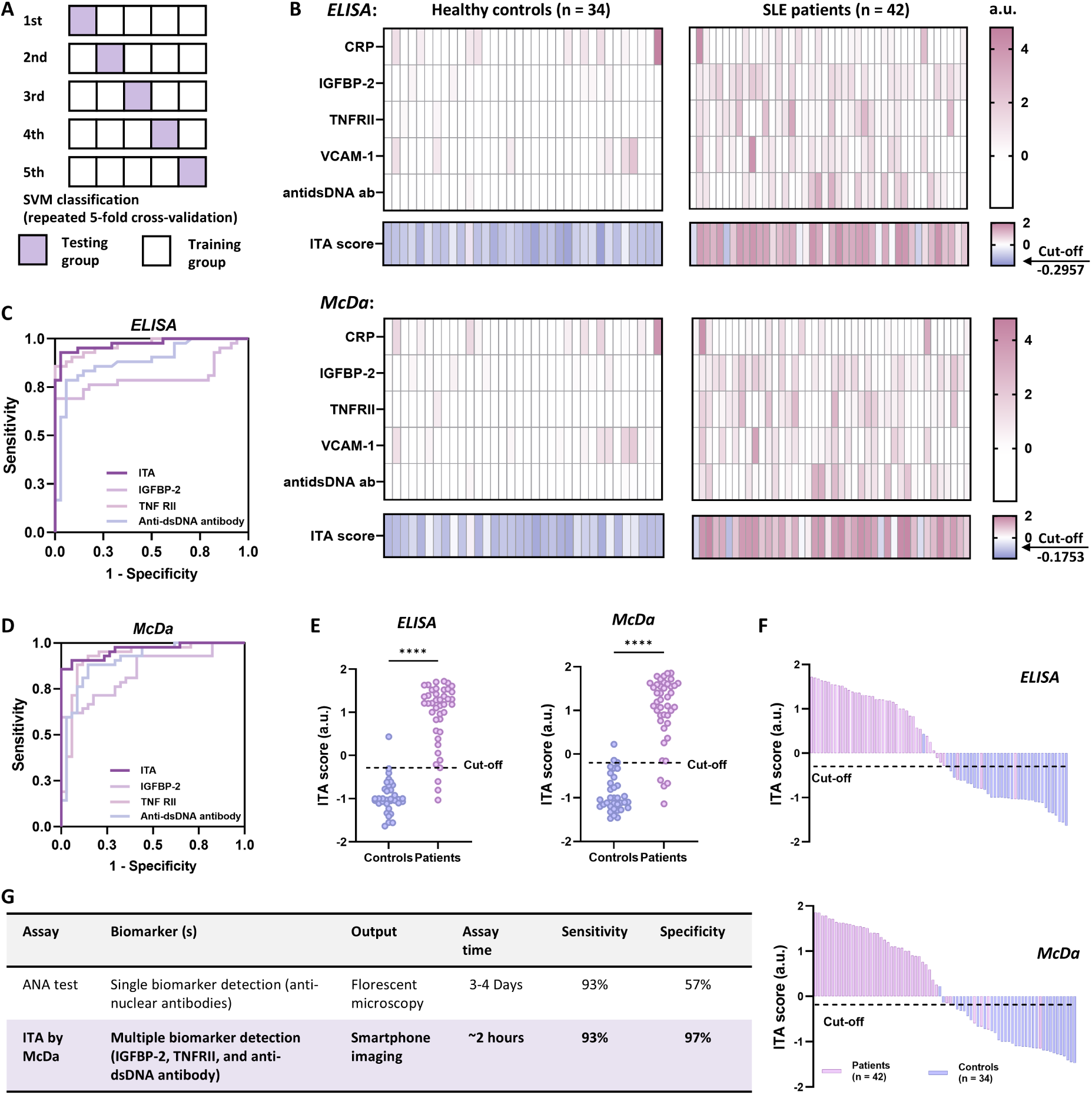
Machine learning to identify an optimal biomarker panel—“ITA” for SLE diagnosis. **A.** Schematic illustration of the five-fold cross-validation. **B**. The heat maps show normalized expressions of five individual biomarkers (CRP, IGFBP2, TNFRII, VCAM-1, and anti-dsDNA antibody) in healthy controls (n=34) and SLE patients (n= 42), measured by either ELISA or McDa. An “ITA” comprising **I**GFBP2, **T**NFRII, and **A**nti-dsDNA antibody was established by a machine learning model of SVM with 5-fold cross-validation (also summarized in Table 1). The ITA scores can distinguish SLE patients from healthy controls with a clear cut-off value of -0.2957 and -0.1753 for ELISA and McDa, respectively (bottom in each panel in **B**). **C&D**. Receiver operating characteristic (ROC) analysis verified the capability of these three biomarkers to discriminate between SLE patients and healthy controls. The ITA index demonstrated the best diagnostic capability over individual biomarkers, with AUC values of 0.9727 and 0.9629, respectively, as measured by either McDa or ELISA. **E**. The cut-off value represents the optimized sum of sensitivity and specificity generated by ROC analysis. The ITA scores in SLE patients were significantly higher than those in controls. **F.** The cut-off value of ITA can differentiate SLE patients from healthy controls. **G**. The diagnostic score ITA by McDaA is compared to ANA test. a.u., arbitrary units. **** P< 0.0001 by two-tailed unpaired Student’s *t*-test.

**Table 1.** The comparison of SLE diagnostic statistics of selected biomarker penal and traditional ANA testing.

| ELISA: |  |  |  |  |  |  |  |  |  |  |  |
| --- | --- | --- | --- | --- | --- | --- | --- | --- | --- | --- | --- |
| Biomarkers |  | Training cohort |  |  |  | Testing cohort |  |  |  | Cut-off | AUC |
|  |  | AUC | Sensitivity (%) | Specificity(%) | Accuracy(%) | AUC | Sensitivity (%) | Specificity(%) | Accuracy(%) |  |  |
| Single | CRP | 0.6455 | 80.61 | 30.56 | 58.58 | 0.4881 | 75.56 | 21.43 | 50.08 | 0.7208 | 0.6380 |
|  | IGFBP-2 | 0.8099 | 69.02 | 91.85 | 79.26 | 0.7855 | 68.61 | 88.57 | 77.58 | 0.3005 | 0.7997 |
|  | TNFRII | 0.8994 | 88.72 | 82.35 | 85.86 | 0.9049 | 88.61 | 82.38 | 85.58 | 0.2026 | 0.8992 |
|  | VCAM-1 | 0.6798 | 71.37 | 54.29 | 63.82 | 0.5455 | 65.56 | 44.29 | 55.25 | 0.4290 | 0.6530 |
|  | Anti-dsDNA antibody | 0.8826 | 72.01 | 94.10 | 81.91 | 0.8650 | 70 | 93.81 | 80.42 | -0.1819 | 0.8852 |
| Panel | TNFRII + anti-dsDNA antibody | 0.9727 | 88.73 | 89.71 | 89.15 | 0.9579 | 90.83 | 88.10 | 89.58 | -0.4063 | 0.9531 |
|  | **IGFBP-2 + TNFRII + anti-dsDNA antibody** | **0.9893** | **92.89** | **98.52** | **95.39** | **0.9790** | **93.33** | **97.14** | **94.75** | **-0.2957** | **0.9727** |
|  | TNFRII + VCAM-1 + anti-dsDNA antibody | 0.9790 | 90.48 | 90.45 | 90.46 | 0.9507 | 90.83 | 84.76 | 88.25 | -0.4760 | 0.8620 |
|  | IGFBP-2 + TNFRII + VCAM-1 + anti-dsDNA antibody | 0.9886 | 92.89 | 97.78 | 95.07 | 0.9612 | 90.83 | 94.29 | 92.17 | -0.08759 | 0.9580 |
|  | CRP + IGFBP-2 + TNFRII + VCAM-1 + anti-dsDNA antibody | 0.9856 | 93.48 | 98.52 | 95.73 | 0.96 | 85.83 | 93.81 | 89.42 | -0.1533 | 0.9524 |
| Traditional test | ANA test |  |  |  |  |  | 93.00 | 57.00 |  |  |  |

| McDa: |  |  |  |  |  |  |  |  |  |  |  |
| --- | --- | --- | --- | --- | --- | --- | --- | --- | --- | --- | --- |
| Biomarkers |  | Training cohort |  |  |  | Testing cohort |  |  |  | Cut-off | AUC |
|  |  | AUC | Sensitivity (%) | Specificity(%) | Accuracy(%) | AUC | Sensitivity (%) | Specificity(%) | Accuracy(%) |  |  |
| Single | CRP | 0.6061 | 78.82 | 32.72 | 58.55 | 0.4792 | 70.28 | 21.43 | 47.50 | 0.9172 | 0.5798 |
|  | IGFBP-2 | 0.8373 | 69.07 | 84.50 | 75.99 | 0.8146 | 63.89 | 77.14 | 69.92 | 0.8526 | 0.8459 |
|  | TNFRII | 0.9261 | 90.50 | 87.49 | 89.15 | 0.9169 | 88.33 | 88.10 | 88.17 | 0.2041 | 0.9230 |
|  | VCAM-1 | 0.6719 | 79.13 | 40.24 | 61.85 | 0.3890 | 67.5 | 24.29 | 47.25 | 0.9922 | 0.6373 |
|  | Anti-dsDNA antibody | 0.9005 | 75.54 | 89.66 | 81.90 | 0.8950 | 71.94 | 87.62 | 79.08 | -0.2502 | 0.9062 |
| Panel | TNFRII + anti-dsDNA antibody | 0.9686 | 92.89 | 83.78 | 88.81 | 0.9587 | 93.33 | 84.76 | 89.50 | -0.3363 | 0.9461 |
|  | **IGFBP-2 + TNFRII + anti-dsDNA antibody** | **0.9836** | **94.67** | **97.06** | **95.73** | **0.9751** | **93.33** | **97.14** | **94.83** | **-0.1753** | **0.9629** |
|  | TNFRII + VCAM-1 + anti-dsDNA antibody | 0.9810 | 94.65 | 92.65 | 93.754 | 0.9410 | 93.33 | 84.76 | 89.50 | -0.6591 | 0.8382 |
|  | IGFBP-2 + TNFRII + VCAM-1 + anti-dsDNA antibody | 0.9902 | 94.67 | 95.58 | 95.08 | 0.9754 | 95.56 | 90.95 | 93.50 | -0.1839 | 0.9433 |
|  | CRP + IGFBP-2 + TNFRII + VCAM-1 + anti-dsDNA antibody | 0.9897 | 93.48 | 93.48 | 96.32 | 0.9716 | 90.56 | 87.62 | 89.50 | -0.1986 | 0.9425 |
| Traditional test | ANA test |  |  |  |  |  | 93.00 | 57.00 |  |  |  |

### Diagnostic specificity improvements by ITA

Importantly, the diagnostic specificity of ITA for SLE diagnosis in the testing cohort was found to be 97.14% compared with 57.00% for the current clinical standard ANA test, representing a 70.42% improvement in specificity, while retaining the sensitivity as the ANA test. Because the specificity of the ANA test is low, it means a higher probability of a false positive diagnosis or overdiagnosis, leading to unnecessary treatments. ITA measurement can markedly improve the diagnosis, as opposed to the current standard of the ANA test. To the best of our knowledge, this SLE diagnostic score powered by machine learning represents the best set of biomarkers for SLE diagnosis, warranting further clinical validation in the future.

### Design, fabrication, and workflow of miniDia for translating McDa into PoC diagnostics

The signal conversion from 3D microneedles to 2D gel made it possible for data acquisition with a smartphone-based App. We thus designed and built a cost-efficient and friendly operative “all-in-one” PoC device for automatically McDa assaying without requiring special training or a laboratory (Fig. 6A&B). The major parts of the prototype were fabricated by 3D printing at a palm-sized 12.5 (L) x 8.5 (W) x 8.3 (H) cm, powered by a rechargeable Li-polymer battery and operated by a microcontroller unit (MCU). It was composed of an immunoassay station (Fig. 6C, blue), a reagent-prefilled millifluidic cartridge (Fig. 6D; Photograph in Supplementary Fig. S12), and an imaging module (Fig. 6C, pink). The immunoassay station contains a vacuum chamber, a waste container, an MNA housing chamber, and a one-way valve (Fig. 6C, blue). A smartphone for taking images can be placed on top of the imaging station that holds a microlens, a ring-LED light (broad-wavelength white light), and a gel station (Fig. 6C, pink).

**Fig. 6.**
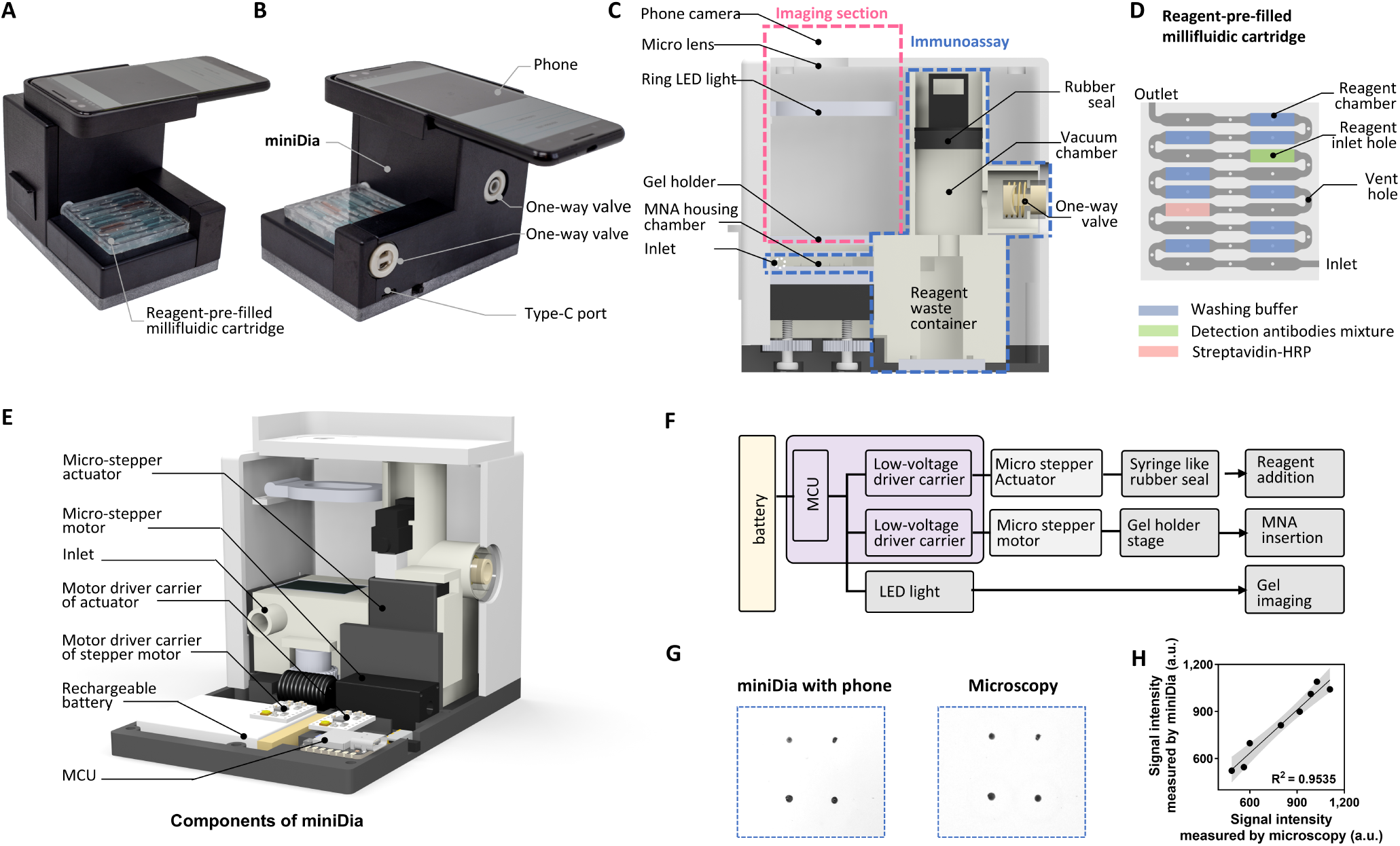
miniDia design and fabrication. **A.-B**. Photographs of the miniDia prototype with a smartphone. **C**. Schematic diagram of a vertical cross-section of the miniDia composed of two main parts, the immunoassay station (blue outline) and the imaging section (pink outline). **D**. Illustration of the reagent-pre-filled millifluidic cartridge with chambers colored for different solutions (Photograph in Supplementary Fig. S12). **E.** illustration of internal architecture of the device, also Photograph in Supplementary Fig. S13. **F.** Block diagram of control panels of the automated miniDia device. **G**. Representative images of the same stained gel acquired by miniDia with a smartphone and microscopy**. H**. The paired correlation test was conducted for the McDa versus the microscopic results. a.u., arbitrary units.

As shown in Fig. 6D, the reagent-prefilled millifluidic cartridge contains 18 segments, one inlet port equipped with a one-way valve for airflow control, and one outlet port connected to the immunoassay station. A sealing film was applied over the inlet holes after reagent loading to protect the solutions during storage and operation and to prevent evaporation. Each reagent segment was sequentially prefilled through an inlet hole, while the vent hole allowed air to escape during reagent filling. Air gaps between the two segments prevented reagent intermixing. To better visualize the reagents within the chambers, solutions were marked in different colors, and a photograph of the cartridge is shown in Supplementary Fig. S12.

The MNA housing chamber is connected to the reagent-prefilled millifluidic cartridge at one end and to a waste container at the other end. After the MNA patch captures target biomarkers and is loaded into the device, the device automatically performs sequential washing, detection-antibody incubation, streptavidin-HRP incubation, and final washing steps within the chamber to form immune-sandwich complexes. Reagent flow is controlled by an “on-demand vacuum” system driven by a micro-stepper actuator, which automatically generates negative pressure to sequentially draw prefilled reagents from the millifluidic cartridge into the immunoassay station (please see Video S2 and Supplementary Fig. S14). For miniDia operation, the user first downloads the miniDia App, registers an account, and logs in, followed by scanning the QR code on the package (Supplementary Fig. S15). The outlet of reagent-prefilled millifluidic cartridge is connected to the inlet of the immunoassay station, while the opposite end is connected to a one-way valve for airflow control (Fig. 6D-E). Then, the user applies an MNA patch pre-coated with capture elements to bind target biomarkers from samples such as blood, saliva, respiratory sweat, or urine. For instance, we have developed a wearable photonic device, termed <u>P</u>hotothermal-induced <u>E</u>xtravasation <u>D</u>evice “PiED” for painless, and minimally invasive sampling of multiple blood biomarkers without blood drawing^38^. Application of PiED to the skin induces biomarker extravasation from skin capillaries via light-transmitted MNA (OMNA), enabling the extravasated biomarkers to bind the surface of OMNA in a one-biomarker-per-microneedle fashion. The MNA patch and gel can be loaded into the miniDia for automatically process (Supplementary Fig. S15 and Video S2). The gel chamber is positioned above the MNA housing and separated by a gel cover, which prevents reagent contamination during the assay while also serving as a white background for imaging (Supplementary Fig. S14).

Finally, the imaging module is integrated into a light-shielded chamber to enable imaging of the stained gel (Fig. 6C, pink section). A second stepper motor, controlled by the same MCU, raises the MNA stage through a gear-train mechanism and inserts the stained MNA into the gel, enabling conversion of the 3D microneedle signals into a 2D format for cellphone imaging. Signal images within the gel are further amplified using a microlens under LED illumination. A ring-LED surrounding the chamber wall provides uniform illumination across the gel and ensures consistent imaging conditions independent of ambient light variability. Representative images captured by miniDia and by microscopy are shown in Fig. 6G. The two methods showed a high correlation, demonstrating the clinical potential of miniDia for PoC applications (Fig. 6H). The total assay time of the McDa-miniDia platform, from sample collection to result readout, is approximately 2 hours.

## Discussion

Translating a disease-specific biomarker panel into PoC or home testing requires more than demonstrating its diagnostic performance; it requires a sensing platform capable of measuring all selected biomarkers reliably under the practical constraints of decentralized testing. Unlike centralized laboratory testing, decentralized testing typically relies on microliter-scale fingerstick samples and a single sample dilution; e.g., biomarkers spanning widely different abundance ranges must be quantified within the linear range of a single assay in a single sample dilution. Moreover, multiple targets must be measured under the same chamber without cross-reactivity, generating reliable and specific signals for each biomarker. Thus, a biomarker panel optimized solely for statistical discrimination may not be practically measurable by a PoC assay, while a sensor optimized solely for analytical sensitivity may not meet the requirements of multiplex clinical testing. Decentralized diagnostics therefore require coordinated optimization of biomarker selection, assay design, multiplex measurement, automated processing, and computational interpretation rather than independent optimization of individual components.

The current study establishes a **bench-to-decentralized diagnostic platform spanning the entire translational pathway,** from disease-specific biomarker panel discovery using clinical samples to the development of a highly sensitive multiplex assay, automated data analysis, and decentralized PoC testing (Fig. 1). At the center of this pathway is McDa, which serves as the analytical interface between an optimized biomarker panel and its practical implementation as a multiplex assay. McDa combines one-biomarker-one-dot multiplex detection with chemo-spatial signal amplification and smartphone-compatible colorimetric readout, while **miniDia** automates the assay and signal acquisition. Together with ML-guided biomarker panel optimization and intelligent image analysis, these components form an integrated workflow from biomarker selection to decentralized diagnostic readout. Importantly, by integrating these components into a single platform, the iPlexD2Go closes the translational gap between biomarker discovery and clinically actionable diagnosis, achieving superior laboratory-grade sensitivity without the need for centralized laboratory infrastructure. When comparing representative microarray and PoC technologies from the literature in terms of sensing modality, an AI-defined disease-specific biomarker panel, clinical diagnostic improvement, home-use potential and associated concerns, and analytical sensitivity (Supplementary Table S4), iPlexD2Go is the only platform that satisfies all of these criteria. Therefore, iPlexD2Go represents an important advance toward the practical implementation of precision diagnostics in decentralized settings.

A versatile and robust McDa colorimetric assay was essential for the highly sensitive multiplex biomarker detection of the iPlexD2Go and offers several advantages. First, conjugating a specific antibody onto a designated microneedle within an MNA enables detection of dozens of biomarkers on a single array in a one-biomarker-per-microneedle format. Second, detection signals are robustly amplified through a multiple-signal enhancement strategy involving surface-area enlargement, sequential chemical surface modifications, volumetric color precipitation, and dimensional signal conversion. Third, converting signals from 3D microneedles into a 2D gel format greatly facilitates cost-effective and efficient smartphone-based data acquisition and analysis. Fourth, the platform can be readily adapted to diagnosis of a broad spectrum of diseases, including chronic diseases, organ rejection, cancers, and acute infectious diseases, using various sample types. Moreover, we identified an optimized three-biomarker panel capable of discriminating patients with SLE from healthy controls. The newly developed disease score (ITA) was generated through deep-learning-assisted modeling and a support vector machine (SVM) model with 5-fold cross-validation, which substantially outperformed the current clinical standard antinuclear antibody (ANA) test. Importantly, these biomarkers can be reliably and sensitively measured by iPlexD2Go and reported and analyzed on a smartphone.

Traditionally, multiple biomarker detection depends heavily on centralized analytical laboratories equipped with costly specialized instrumentation and trained healthcare workers to conduct time-consuming assays. Patients with chronic diseases, organ transplants, or undergoing cancer therapies often require close and continuous monitoring of disease progression and therapeutic responses. The availability of a PoC device capable of measuring multiple biomarkers at home would allow patients to perform the tests without frequent hospital visits. Such self-tests could save time, reduce healthcare and transportation costs, enable timely intervention, and lower the risk of disease exacerbation, which are increasingly important with the rapid expansion of digital health and home-based care. Currently, most reported MNA-based biosensors are limited to the detection of a single biomarker per MNA^39,40^. For instance, Wang et al. reported an MNA assay using a gold nanorod-enhanced fluorescent labeling for on-microneedle detection of a single biomarker with high sensitivity; however, the signal acquisition and analysis relied on confocal microscopy^40^. Achieving multiplex biomarker detection using microneedle arrays is particularly challenging because the miniature architecture of each microneedle makes selective functionalization with distinct capture antibodies on individual microneedles technically demanding. In addition, immunosignals are generated on the three-dimensional surfaces of individual microneedles, requiring confocal microscopy for one-by-one image acquisition and analysis. Although several studies have reported MNAs for multiplex biomarker detection, most achieved multiplexing through regionally separated sensing electrodes or sensing zones, and none have demonstrated the ability to immobilize a specific capture element on a designed microneedle within a single MNA^22,41^. For example, Tehrani et al. fabricated a wearable microneedle array for real-time monitoring of multiple metabolites, but the multiplexing was achieved by using microneedle-electrode regions functionalized with analyte-specific enzyme layers for different biomarkers, rather than functionalizing individual microneedles within the same array^22^. In marked contrast, the mask-assistant strategy developed in this study allows covalent conjugation of different capture antibodies on individual microneedles, representing, to the best of our knowledge, the first approach for multiple biomarker measurement with a single MNA via microneedle-surface functionalization.

Apart from enabling the conjugation of a specific capture antibody to a designed microneedle within the MNA, the MNA serves two essential functions in the McDa platform. First, its 3D architecture substantially increases the effective detection surface area compared to a 2D flat dot array, enhancing analyte capture efficiency. Second, and more importantly, the MNA can penetrate a substrate-saturated gel, enabling the conversion of 3D immunosignals into a 2D colorimetric readout. Another key contributor to the exceptional sensitivity of McDa is the dendron decoration of the MNA surface, which effectively overcomes the limited number of reactive sites achievable with oxygen plasma treatment and PEI modification alone^42,43^. A myriad of primary amine groups provided by the dendron dramatically increases the number of activated conjugation sites for capture antibody immobilization on each microneedle, which alone increases the signal intensity by 28-fold^44^. These structural and surface-engineering innovations endow the McDa platform with exceptional analytical sensitivity. Another major challenge in multiplex biomarker detection is accommodating biomarkers with vastly different physiological concentrations within a single assay. Unlike conventional ELISA, where each biomarker can be measured using an independently optimized sample dilution, multiplex assays require a single serum dilution for the simultaneous quantification of all biomarkers. For example, a low-abundance biomarker such as TNFRII may require minimal serum dilution (e.g., 1:50) for accurate detection, whereas a highly abundant biomarker such as CRP may require dilution greater than 1:5,000 to remain within the assay’s linear range. Reconciling these disparate concentration ranges within a single measurement presents a significant analytical challenge. In this regard, McDa exhibited wide linear ranges and low LoD values comparable to those of traditional ELISA. Moreover, the density of capture antibodies on individual microneedles was optimized according to the expected abundance of each target biomarker, with lower-abundance biomarkers assigned higher capture antibody densities and higher-abundance biomarkers assigned lower densities. This design enabled the simultaneous quantification of all biomarkers using a single serum dilution while maintaining analytical accuracy across a wide concentration range. Finally, while signal quantification in conventional laboratory assays is typically performed on specialized instrumentation and computer software, PoC testing requires automated analysis and user-friendly result interpretation. To address this challenge, we trained a deep-learning model using manually annotated gel images and integrated it into smartphone-based quantitative analysis, enabling automated biomarker quantification and user-friendly result reporting.

SLE is a chronic autoimmune disease manifested with various abnormalities in multiple organs, which can eventually lead to tissue and organ damage if left uncontrolled^45,46^. Continuous disease activity monitoring is critical for effective therapy and disease management to minimize the risk of organ damage and reduce mortality. Because of the increased presence of autoantibodies in SLE, a positive ANA test is the gold standard in the 2019 SLE classification criteria^47,48^. The indirect immunofluorescent assay is carried out by skilled lab technicians with cell culture and a fluorescent microscope in a well-equipped lab,^48^ and it takes days for the result report. Even with this sophisticated laboratory testing, the specificity is only 57%, leading to a high possibility of false positives and overtreatment, greatly complicating disease management. This well exemplifies how a single biomarker is often insufficient for accurate disease diagnostics^27^. On the contrary, the ITA score, defined in the current study by leveraging the machine learning model using SVM combined with a 5-fold cross-validation of clinical samples, shows great potential for SLE diagnosis and monitoring. In this machine learning modeling, we randomly split the samples into five groups and employed 5-fold cross-validation to assess the biomarker’s diagnostic potential individually. The AUCs for IGFBP-2, TNFRII, and anti-dsDNA antibody ranked the highest among the five biomarkers, consistent with previous demonstration of various biomolecules’ involvement in SLE development, besides autoantibodies^49–51^. The three (ITA) biomarkers reflect different pathways of immune cell proliferation and apoptosis (IGFBP-2), B-cell activation and related production of autoantibodies (TNFRII), and autoantibodies targeting ds-DNA (anti-dsDNA), thus conferring a better diagnosis, compared to a single biomarker (Table 1). Compared with the ANA test, McDa for measuring the ITA score is convenient for at-home self-testing, cost-effective, and offers higher specificity (97%) with comparable sensitivity (93%) (Fig. 5G). Limitations of the study include a relatively small sample size, which needs to be expanded to include a larger, more diverse cohort from multiple clinical sites to validate the potential of the McDa before establishing the ITA as a standard for SLE diagnosis.

In summary, the “iPlexD2Go” provides femtomolar sensitivity and specificity for measuring a panel of biomarkers with only <0.1 µL of the sample in a rapid, cost-effective, and self-testing manner without the need for a laboratory or professional personnel. It measures both low- and high-abundance biomarkers in a single assay at a single dilution. By leveraging machine learning, we also identified a biomarker panel, containing IGFBP-2, TNFRII, and anti-dsDNA antibodies, which performed best in distinguishing SLE patients from healthy controls and can be directly measured by iPlexD2Go. Thus, iPlexD2Go provides an end-to-end systems-level framework for translating disease-specific biomarker panels into deployable diagnostics for decentralized diagnosis, enabling simple, precise, and efficient data acquisition and analysis, making it easily adaptable to PoC or home use for diagnosis and monitoring of many other diseases, in addition to SLE.

## Materials and Methods

### Materials

Poly(methyl methacrylate) (PMMA) (mw∼120,000), poly(ethyleneimine) (PEI) (M_n_∼60,000; M_w_∼750,000), ethyl acetate, polyamidoamine (PAMAM) dendrimer (cystamine core, generations 2.0, 4.0, and 5.0), tris(2-carboxyethyl)phosphine hydrochloride (TCEP), dihydrazide (AAD), alginate, 1-Ethyl-3-(3-dimethylaminopropyl)carbodiimide (EDC), *N*-hydroxysuccinimide (NHS), suberic acid bis(3-sulfo-N-hydroxysuccinimide ester), sodium hydroxide (NaOH), isopropanol, double-stranded DNA (dsDNA), albumin methylated from bovine serum (mBSA), and bovine serum albumin (BSA) were all purchased from Sigma-Aldrich (Waltham, MA, USA). SeramunBlau spot dark 3,3’,5,5’-Tetramethylbenzidine (TMB) substrate was obtained from Seramun Diagnostica GmbH (Heidesee, Germany). Biotin-Goat Anti-Human IgG was purchased from Jackson ImmunoResearch (West Grove, PA, USA). Streptavidin-horseradish peroxidase (HRP) was purchased from Abcam (Waltham, MA, USA). Polydimethylsiloxane silicone (PDMS) elastomers base and curing agent (SYLGAR 184 Silicone Elastomer Kit) were obtained from Dow (Midland, MI, USA). PBS was purchased from Life Technologies (Carlsbad, CA, USA). MES buffer was obtained from Thermo Scientific (Rockford, IL, USA). Human C-reactive protein (CRP) (DY1707), Human insulin-like growth factor binding protein-2 (IGFBP-2) (DY674), Human sTNF RII/TNFRSF1B (DY726), and Human VCAM-1/CD106 DuoSet ELISA kits and sealing films were purchased from R&D System (DY809) (R&D Systems, Minneapolis, MN, USA). TPU adhesive patch was obtained from 3M (Maplewood, MN, USA). Distilled water was obtained from a Millipore Milli-Q ultrapure water purification system (Burlington, MA, USA).

### Clinical samples

We obtained serum samples from lupus patients (N = 42) and healthy controls (N = 34) from Mass General Brigham Biobank, the University of Houston (UH), and University of Texas Southwestern Medical Center (UT Southwestern). The samples from UH and UT Southwestern were following the UH approved IRB protocols. Consent was obtained from all subjects before sample collection. All samples were aliquoted and stored at –80 °C until use.

### Fabrication of PDMS-MNA mold

An MNA was fabricated according to the previous study with some modifications^52^. Briefly, an MNA mold fabricated in our previous study was used for the fabrication of PDMS-MNA mold^52^. PDMS elastomer base solution mixed with curing agent at a 10:1 ratio was poured into a well of a 6-well plate, and mixed well, followed by centrifuge at 2,000 rpm for 10 min to remove bubbles. The original MNA mold was then placed into the mixture, and bubbles were removed under a vacuum. The mixture was then heated at 85°C for 3 hours. The original MNA mold was removed after cooling to obtain a female PDMS-MNA mold. PMMA solution at 1 mL was added into the female PDMS-MNA mold, followed by centrifugation at 4,000 rpm for 15 min.

### Fabrication of MNA and surface modifications

The PMMA solution (20% w/v) was prepared by dissolving at ethyl acetate and stirring at 78°C overnight before its addition to the female PDMS-MNA mold. The casting process occurred at 80 °C for 4 h to remove ethyl acetate, followed by adding 1 mL of PMMA solution to cover the first layer of dried PMMA at 85 °C overnight. The process was repeated once.

After PMMA was dried and cooled, the resultant MNA was carefully peeled from the female PDMS-MNA mold and washed with 2-propanol three times, followed by air-dry and oxygen plasma treatment with a Technics 500-II Plasma etcher (200 W) for 3 min. The plasma-treated MNA was then immediately immersed in PEI solution (10% v/v, pH 11) at 60 °C and stirred for 6 hours. To obtain PAMAM dendron, PAMAM dendrimer (3 mM) with cystamine core was cleaved by incubation with TCEP (5 mM) for 1 h at RT, followed by removal of TCEP using a desalting column. PAMAM dendron (2 uM) was then conjugated onto the PEI-modified surface of each microneedle on the MNA by using suberic acid bis(3-sulfo-N-hydroxysuccinimide ester) sodium salt (2 uM) as a cross-linking reagent. The surface morphology was characterized by SEM (Hitachi SU5000 VP FE-SEM).

### Immobilization of multiplex capture elements on a single MNA in one capture-element-on-one MN fashion

An array of tiny holes with a diameter of 210 μm precisely aligned with the microneedles on MNA was designed by Solidworks. We used a laser to punch an array of tiny holes on a 330 μm-thick TPU film with a high-strength acrylic adhesive to obtain a mask. The mask was then used to cover the MNA base and each hole formed in the mask served as a STC micro-reaction site for the specific capture antibody immobilization on a designated microneedle in the MNA (Supplementary Fig. S3A). Next, EDC (1 mM) and NHS (2.5 mM) were dissolved in MES buffer (0.1 M), followed by the addition of a capture antibody for 15 min. The capture antibody concentration was inversely related to the abundance of a specific biomarker and pre-defined with pilot tests. After removal of unreactive EDC and NHS by centrifuge filtration, 1 µL of the capture antibody/reaction mixture was pipetted into the designated microneedle to covalently conjugate the antibody via EDC/NHS coupling reaction for 5 h. After careful removal of the mask, the MNA was washed with washing buffer (0.05% Tween-20 in PBS) for three times each 5 min to remove unreacted reagents. Finally, non-specific binding on the MNA was blocked by 2% skimmed milk at 36°C for 1 h and then washed as above. For the detection of anti-ds-DNA antibody, the designated microneedle was incubated with mBSA solution and then ds-DNA solution. The steps of washing and blocking non-specific binding were the same as above.

### Gel preparation

Alginate was dissolved in MES buffer (0.1M) to prepare a 2% w/v solution, followed by the addition of AAD, EDC, and NHS to final concentrations of 10 mM, 40 mM, and 40 mM, respectively. The mixture was cast into a mold to form a hydrogel at room temperature via EDC/NHS mediated AAD crosslinking of alginate carboxyl groups. The resulting hydrogels were thoroughly washed to remove residual reagents. The gels were then cut into 1.4 X 2.1 cm pieces and air-dried, followed by immersion in the SeramunBlau Spot Dark substrate solution for 1 hour at 4 °C in the dark. For long-term storage, the dried hydrogels are vacuum-sealed in bags and stored at 4 °C.

### McDa evaluations

To evaluate the sensitivity and selectivity of McDa, various concentrations of human CRP, mouse CRP, IGFBP-2, TNFRII, VCAM-1, or bovine serum albumin (BSA) solutions were prepared in PBS solution containing 2% BSA, and then added onto designated MNs in the MNA as described above, followed by a 2-h incubation at room temperature (RT). Mouse CRP and BSA were used as negative controls to examine the specificity of the assay. Next, the MNA was rinsed with washing butter every 5 min for a total of three times to remove the non-specific binding. The biotinylated detection antibodies for CRP, IGFBP-2, TNFRII, or VCAM-1 were added to the MNA sequentially and incubated for I hour, followed by washing with washing butter every 5 min for a total of three times and incubation with streptavidin-HRP for 20 min. The MNA was then washed every 5 min for a total of three times. A gel was tailored to a desired size, and air-dried. The dried gel was then immersed in a colorimetric substrate solution until saturated. The MNA was inserted into the gel and pressed firmly for 10 min, after which the gel was removed and washed using distilled water and carefully transferred to a plastic dish for imaging. The images of stained gel were captured by a microscope and analyzed by ImageJ. For single biomarker measurement, the serum samples were diluted 1,000 times for measuring IGFBP-2, TNFRII, and VCAM-1 levels, while serum samples were diluted 20,000 or 40,000 times for CRP detection. For multi-biomarkers measurement in a single MNA, serum samples at 1:1,000 in a reagent diluent were added into the MNs with 2-h incubation at RT. The gel, MNA, and M-chips were imaged by digital microscope (Hayear digital microscope HY2307) or phone camera (Google pixel 3).

### ELISA

CRP, IGFBP-2, TNF-RII, and VCAM-1 were measured sera collected from SLE patients and healthy controls using commercial ELISA kits per the manufacturer’s instructions. To measure anti-dsDNA antibody, a 96-well plate was pre-coated with 0.1 mg/mL of mBSA and incubated for 30 min at 37°C, followed by washing with PBS two times. dsDNA at 200 ug/mL was then added to the plate at 100 uL/well and incubated for 30 min at 37°C, followed by washing as above. The serum samples were diluted 100 times and incubated for 2 h. After incubation with a biotinylated human IgG antibody (2 h) and streptavidin-HRP (20 min) at RT, TMB substrate was added for 20 min. The reaction was stopped by the addition of a stop solution. The results were read on a UV spectrophotometer (Epoch, Biotek, Winooski, VT, USA). The serum samples at a dilution of 1:100 in reagent diluent were used for IGFBP-2, and TNFRII, or at I:1,600 for VCAM-1 detection. The samples at different dilutions of 1:500, 1:10,000, and 1:50,000 in reagent diluent were utilized to quantify CRP.

### Spot recognition by deep learning

We adopted a deep learning-based approach for spot boundary detection using U-Net architecture composed of a “contracting path” and an “expansive path” as previously described.^37^ A total of 1000 spot images were employed as a training group, and 200 spot images as a testing group. The contracting path followed the architecture of a generic convolutional neural network (CNN). It possessed three blocks of double convolution layers, each followed by a rectified linear unit (ReLU) activation function and a 2×2 max pooling operation for downsampling. The contracting served as an encoder, which progressively extracted contextual features while reducing the spatial resolution and increasing the number of feature channels. The expansive path then reconstructed the segmentation map by progressively upsampling the feature maps back to the original image resolution. It consisted of two blocks of up-convolution followed by double convolution layers and ReLU activations. At each upsampling stage, the upsampled feature map was concatenated with the corresponding feature map from the contracting path through skip connections to improve localization accuracy. After the segmentation of spot from the input image, the spot contour was detected and drawn with OpenCV package.

### Design and fabrication of portable prototype miniDia and App development

Apart from the electronic components, micro-stepper actuator, micro-stepper motor, drivers, MCU, rubbers, one-way valves, warm gears, gears, LED, rechargeable battery, and optic lens, all the other components of the miniDia were designed by utilizing a Solidworks 2022 and 3ds Max 2022, converted to STL formation, and printed by 3D printers (including Formlabs Form 4, and Stratasys Objet30). Commercially available rubbers were first tested for their durability and integrated into the miniDia device.

We employed Android Studio (4.0.1) to develop an App using Kotlin as the coding language. The virtual device was Google Pixel 3 (API 30), and the system was Android 10.0+.

### Statistical Analysis

We used the mean signal intensity of a blank plus 3 times the standard deviation of the blank (3σ) to calculate the corresponding biomarker concentration as LoD in Excel. The statistic difference between the two groups was analyzed by a two-tailed unpaired Student’s *t*-test. All graphs were plotted by GraphPad Prism version 9.0. All statistical tests were conducted using GraphPad Prism version 9.0. A p value <0.05 was considered statistically significant for all tests.

To assess the diagnostic ability of selected biomarkers and their combinations, a support vector machine (SVM) model was employed to classify SLE patients from healthy controls using single or combinations of up to five biomarkers. Biomarker values were normalized based on their mean and standard deviation. To prevent overfitting and ensure generalizability, a five-fold cross-validation method was implemented using the scikit-learn package in Python 3.12. Five-fold cross-validation was performed starting with randomly dividing the samples into five groups. Four of the five groups were used as a training set to establish a model, while the remaining group served as the validation set. This process was repeated once, and classification performance metrics, including area under the curve (AUC), accuracy, sensitivity, and specificity, were reported as the average of all five folds during cross-validation for both training and testing sets (Table 1). Subsequently, a final SVM model was constructed using all the data to generate SVM scores, receiver operating characteristic (ROC) curves, as well as to calculate the AUC and cutoff values which were defined as the optimal sum of sensitivity and specificity obtained from ROC curves.

## Acknowledges

This work was supported, in part, by the Defense/Air Force Office of Scientific Research, Military Medical Photonics Program under award number FA9550-17-1-0277, FA9550-20-1-0063, FA9550-23-1-0656, and Department discretionary funds to M.X.W, and the National Institutes of Health (NIH) R01AG062987 to T.W. We also extend our gratitude to Calixto Saenz and The Microfluidics Core Facility at Harvard Medical School for their support in plasma treatment.

## Contributions

M.X.W., T.W., and Z.T.L. conceived the study and designed the research. Z.T.L. conducted experiments. Z.T.L., Y. K. and W.S. prepared and modified the MNA. Z.T.L., P.Y., Y. Z., S.G., Y.L., Y. K., C.L., Z.W., C.T., and M.X.W. analyzed data. C.T., Z.T.L., and S.G carried out ELISA. Z.T.L., P.Y., Y. Z., and W.S. performed bioinformatics analysis and machine learning modeling. W.S., Y.Z. and W.M. contributed to laser treatment on mask and spot segmentation using deep learning. Z.T.L., N.J., S.Z, S.K., B.J., Y.L., W.S., Q.Z., and C.L.E. contributed to the design, fabrication, and photography of the small portable device. Z.T.L. contributed to the 3D animations. Z.T.L. and M.X.W. wrote the paper. T.W. and M.X.W. supervised the project and secured the funding for the study.

## Data availability

All data supporting the results in this study are displayed within the paper and the Supplementary Information.

## Code availability

The code utilized within this paper is available from the corresponding authors upon reasonable request.

