## Supplemental information for "iPlexD2Go Integrates AI-Guided Biomarker Panel Discovery with Decentralized Multiplex Diagnostics"

**Contents**

Supplementary Figures S1–15

Supplementary Tables S1–4

Supplementary Video S1-2

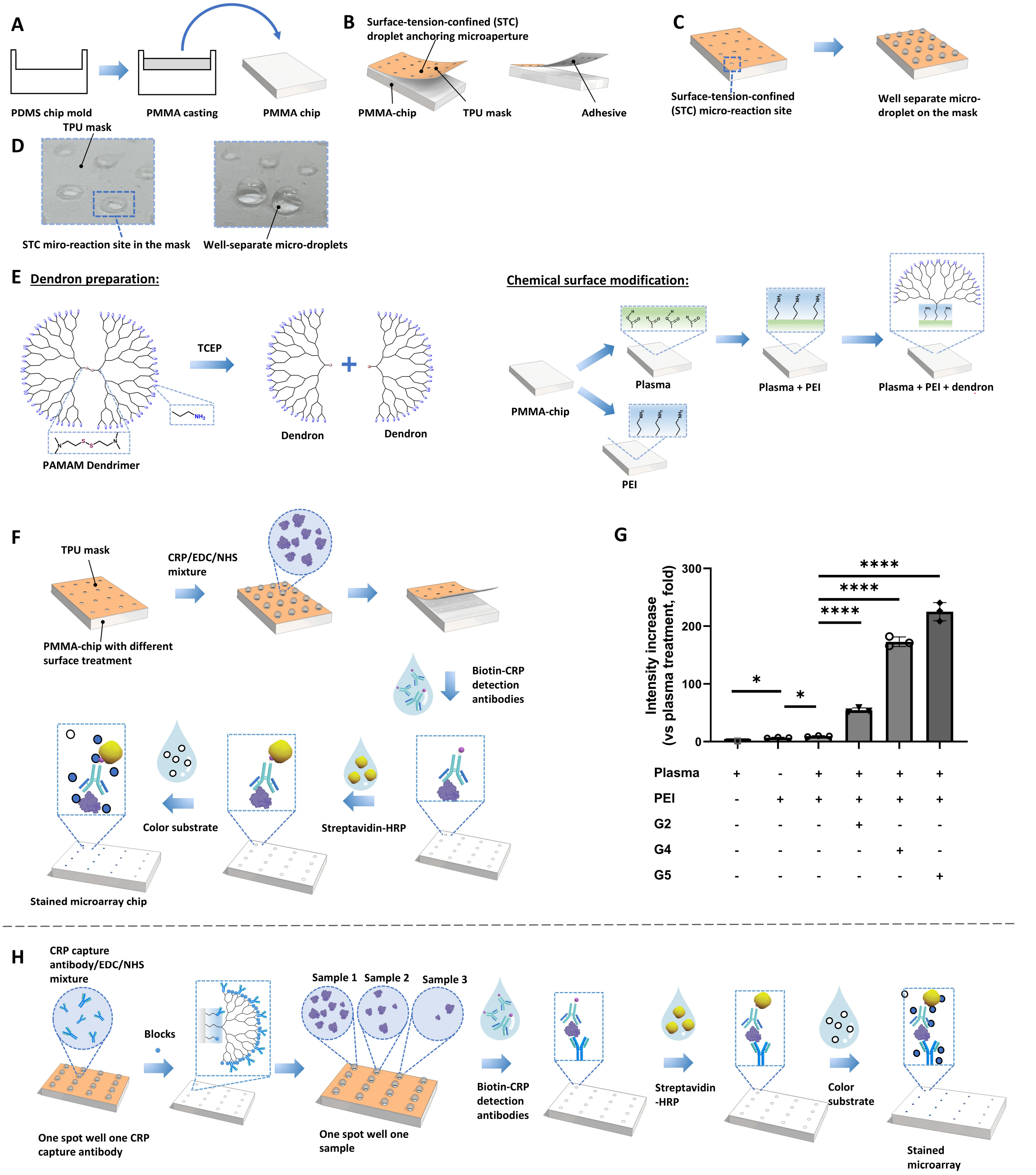


**Supplementary Fig. S1. Construction of PMMA-chip, impact of surface modification on signal intensity, and protein microarray chip**. **A**. Construction of PMMA-chip. **B.** Design of Mask (orange). **C-D**. Illustration (**C**) and photo (**D**) of surface-tension-confined (STC) micro-reaction sites created by a laser-perforated mask adherent on PMMA-chip (top), and solution can be held as STC microdroplet in and above each STC micro-reaction site owing to the hydrophobic surface of the mask (bottom). **E.** Surface modifications of PMMA chip. **F**. Workflow for evaluating signal intensity of antigen-antibody immunocomplex, and photographs of immobilization of multiple capture elements on a single PMMA chip in one capture element on one spot fashion. **G**. Comparison of signal intensity using PMMA chips with different surface modifications. **H.** Construction of protein microarray chip and process of the microarray assay.

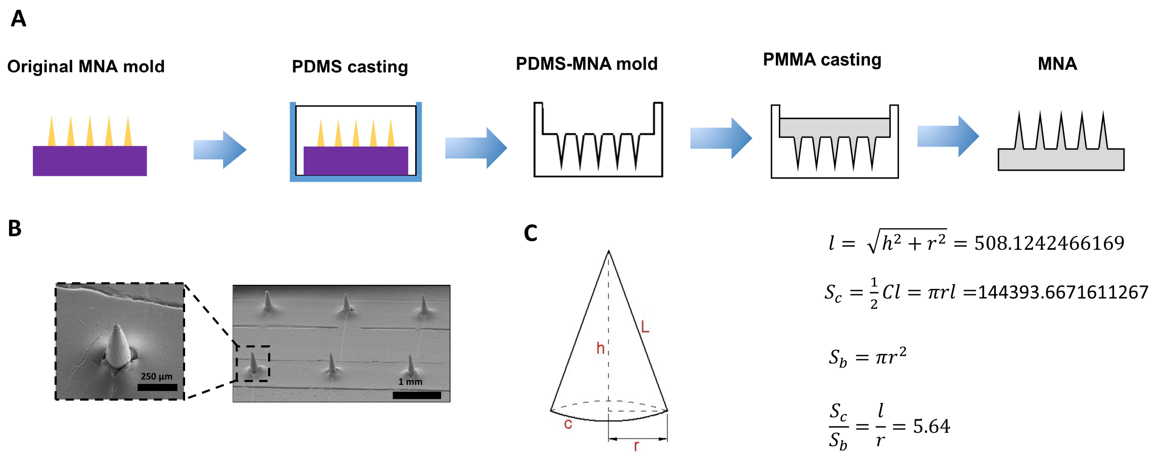


**Supplementary Fig. S2**. **Schematic of fabrication of MNA and SEM image**. **A.** PDMS and cure solution were poured into a well of a 6-well plate, followed by inserting the original male MNA. After polymerization, the PDMS-MNA mold was obtained by removal of the male MNA. The MNA was prepared by casting PMMA solution into the PDMS-MNA mold. **B.** SEM image of the resultant MNA. **C**. The calculation of the ratio between the surface area and the base area of each microneedle.

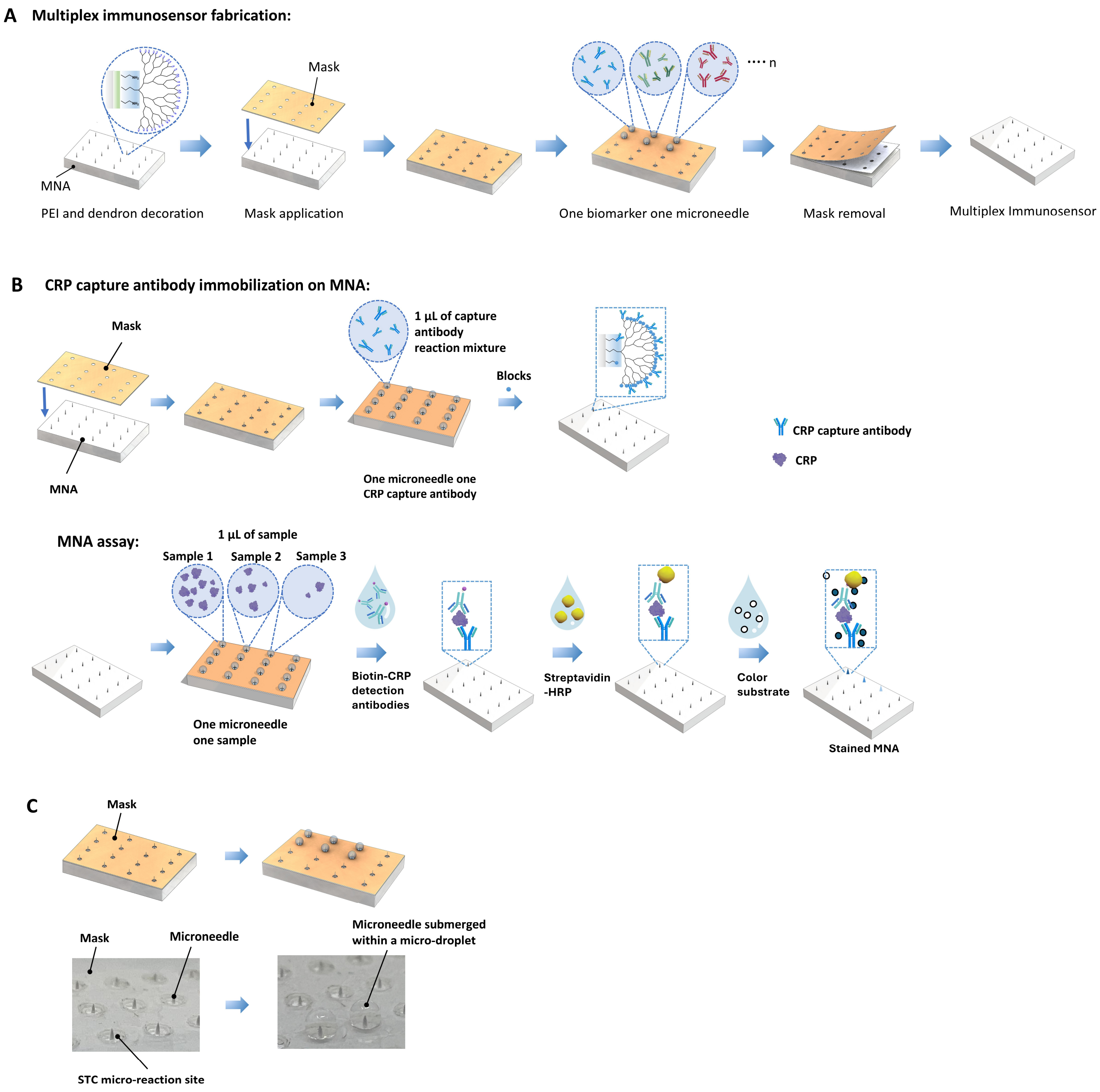


**Supplementary Fig. S3. Illustration of MNA multiplex immunosensor fabrication. A**. Schematic illustration of the immunosensor fabrication and the method for immobilizing multiple capture elements on a single MNA in a one-capture element per microneedle fashion. The MNA surface was modified with PEI and conjugated with PAMAM dendron to exponentially enrich amine groups for capture antibody conjugation. The MNA was then covered by a mask with an aperture pattern precisely aligning with the microneedles in the MNA. These apertures each serve as a STC micro-reaction site to retain an isolated reaction droplet around a single microneedle, thereby allowing the immobilization of a specific capture antibody on a designated microneedle. **B.** Schematic of MNA assay for CRP detection in ultralow volume (1 µL) in one sample on one microneedle fashion. **A**. Illustrations (top) and corresponding photographs (bottom) of immobilizing multiple capture elements on a single MNA in one capture element on one microneedle fashion. Note, individual microneedles are submerged within individual micro-droplets.

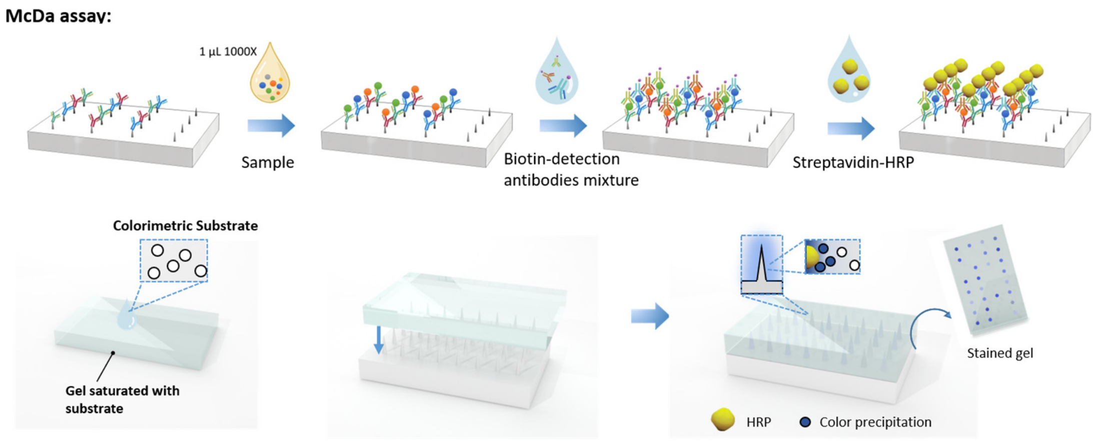


**Supplementary Fig. S4. A. Illustration of McDa.** After washing and immunoassaying, the MNA was inserted into the gel (ice green). The HRP of the immunocomplex catalyzes the substrate within the gel, achieving 3D microneedle sign to 2D gel signal conversion.

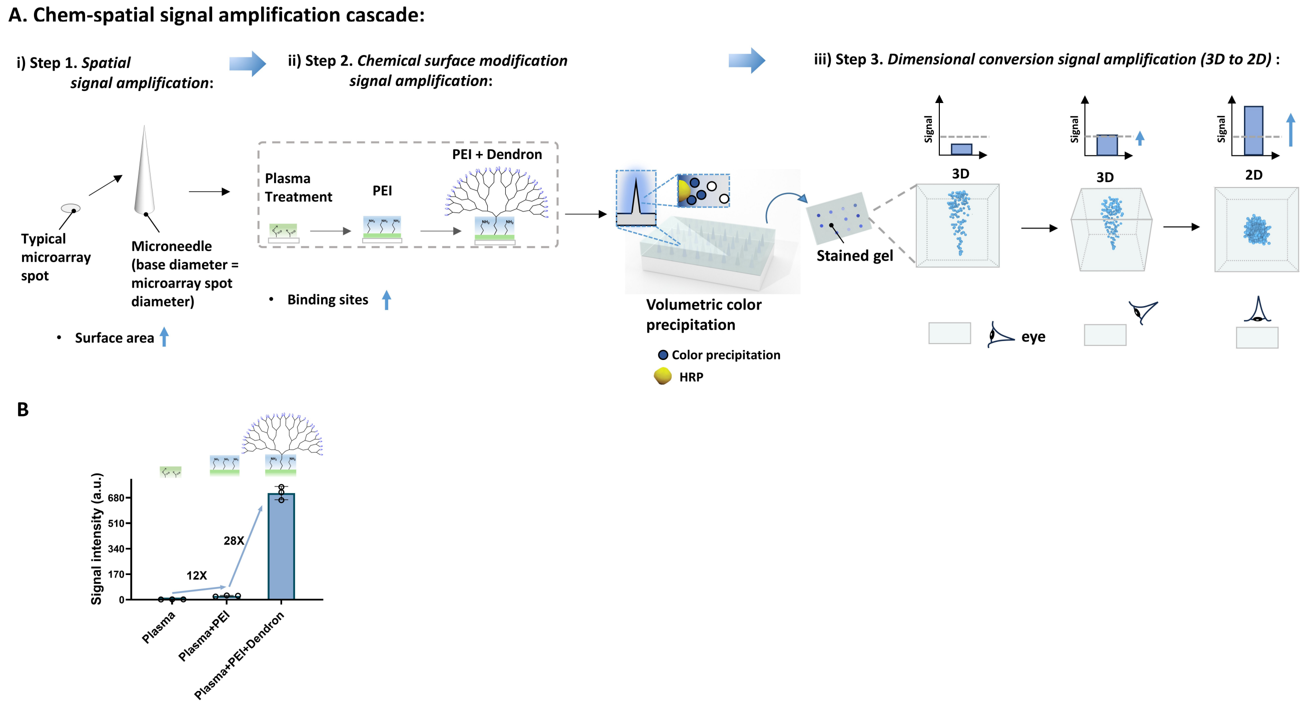


**Supplementary Fig. S5. A. Illustration of chem-spatial signal amplification cascade**, which contains three steps: Introduction of MNA enlarges the surface area; (i). PEI coating and dendron decoration on MNA surface maximize activated binding sites for capture elements, and the 3D microneedle resulted in a volumetric color precipitation distribution in spatial 3D dimensions; (ii). Dimensional conversion from 3D to 2D signal amplification (iii). The top view of the gel surface can be used to observe the maximum color precipitate accumulation. **B.** The signal intensity is amplified by modifying the MNA surface with PEI and dendron (half of dendrimer G5).

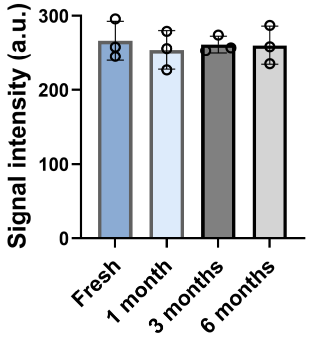


**Supplementary Fig. S6.** The signal intensity for detecting 300 pg/mL CRP by McDa using a substrate-saturated gel stored for 1-6 months.

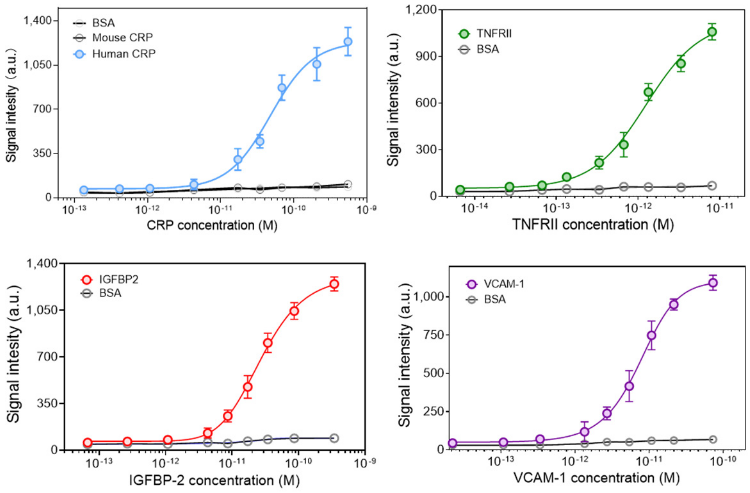


**Supplementary Fig. S7**. Kinetics of McDa responses to an increasing concentration of human CPR, TNFRII, IGFBP-2, VCAm-1, mouse CRP, or BSA in PBS.

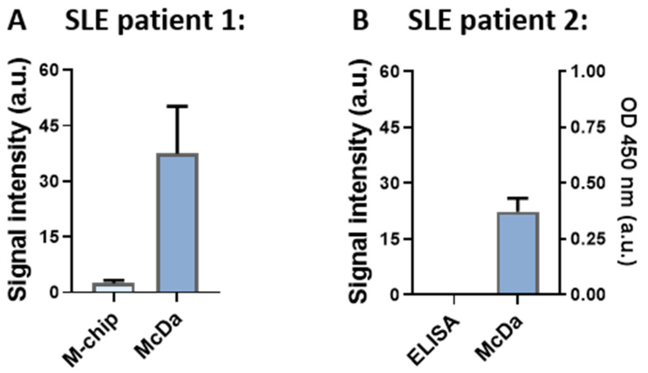


**Supplementary Fig. S8**. The intensity of signal comparison on the same patient sample measured by indicated assays at a dilution of 1:1,000 for TNFRII detection. **A**. For patient 1, the concentration is 5.45 ng/mL. After 1:1,000 dilution, the microarray-chip assay showed no detectable signal due to the low sensitivity of the assay, whereas 5.45 pg/mL was detected by McDa**. B**. For patient 2, after the same dilution, TNFRII was detected by McDa at 3.14 pg/mL, which is lower than the LoD of ELISA. Thus, ELISA failed to detect it.

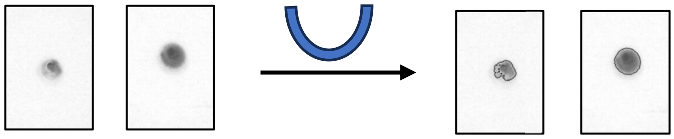

**Supplementary Fig. S9**. **Spot contour recognition**. A deep learning-based approach based on u-net architecture for spot contour recognition on the gel. Shown in S9 are the original spot images (left), a simplified block diagram of u-net architecture (middle), and spot images after contour recognition (right).

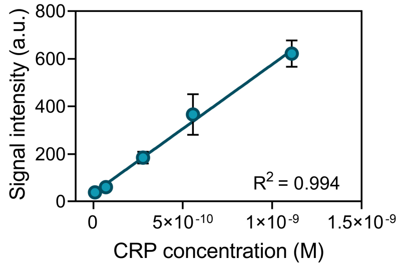


**Supplementary Fig. S10**. Calibration curve of MaDa with multiplex detection for CRP detection.

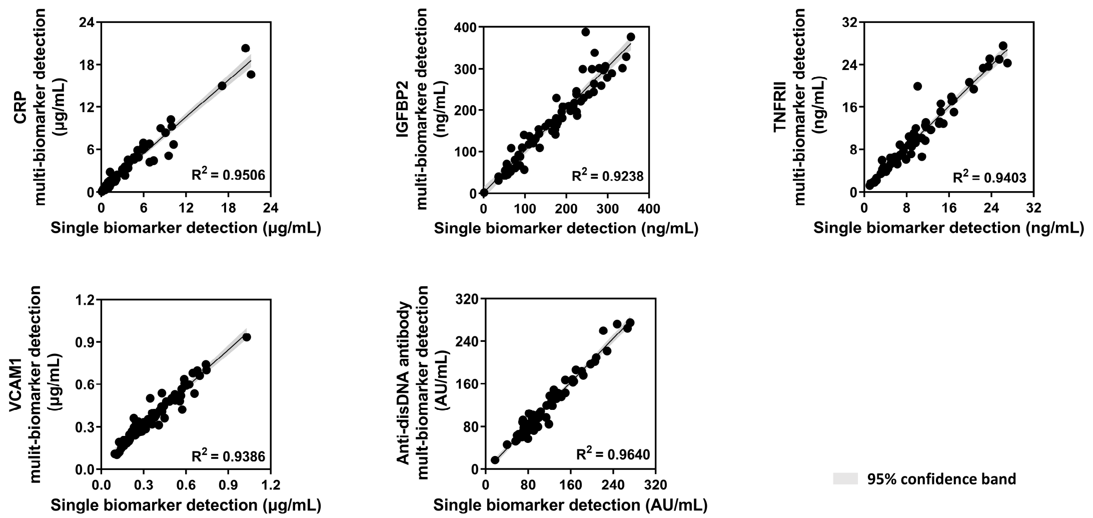


**Supplementary Fig. S11**. The paired correlation tests for single versus multi-biomarker detection by McDa.

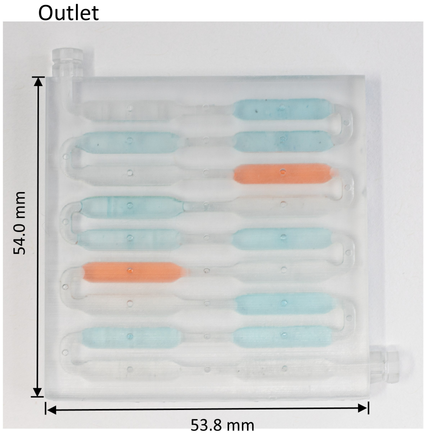


**Supplementary Fig. S12.** Photograph of a reagent-pre-filled millifluidic cartridge with the chambers colored.

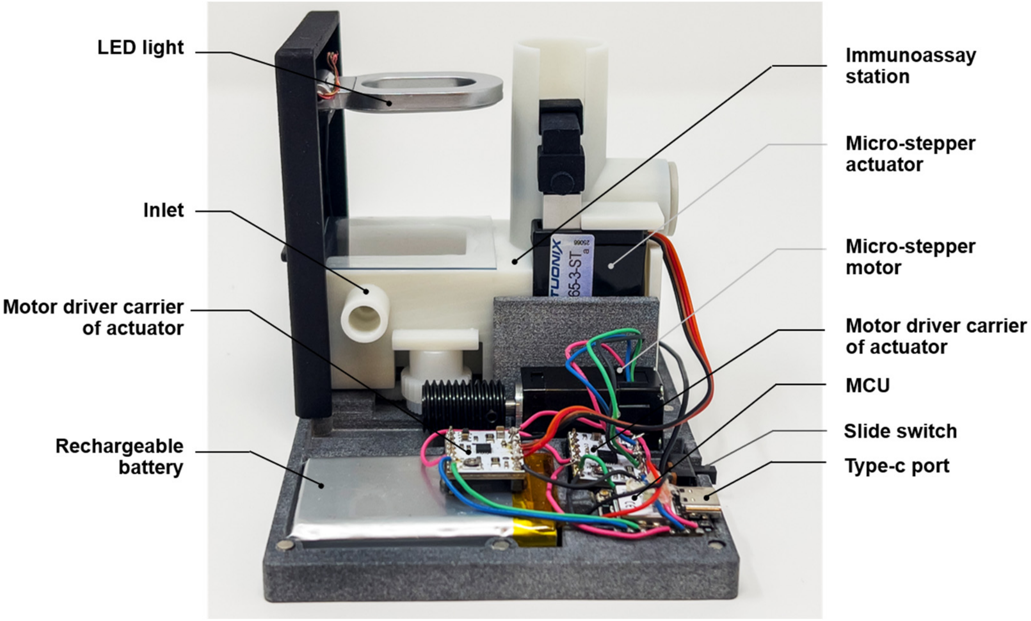


**Supplementary Fig. S13.** Photograph of the internal architecture of miniDia.

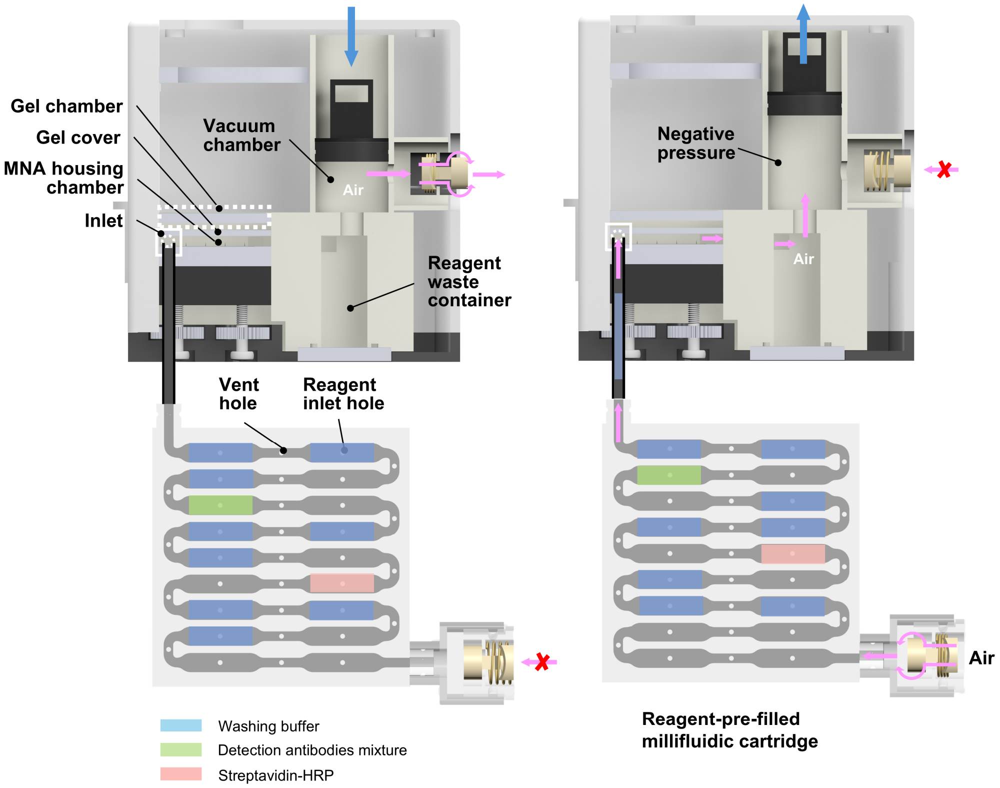


**Supplementary Fig. S14. Illustration of the working principle of "on-demand vacuum" for sequentially drawing the pre-filled reagents in the millifluidic cartridge into the immunoassay station for assaying.** When the rubber seal moves downward, air is expelled from the immunoassay station, waste container, and millifluidic chip through the one-way valve connected to the immunoassay station, while the valve connected to the reagent chip blocks air entry (left). As a result, reagents remain stationary within the chip. When the rubber seal moves upward, the valve connected to the immunoassay station prevents air entry, whereas the valve connected to the reagent chip allows air inflow (right). This creates negative pressure that sequentially pulls the prefilled reagents into the immunoassay station and then into the waste container. The MCU-controlled program drives the micro-stepper actuator according to a preset timing sequence, thereby automating the washing and incubation steps required for immunosandwich formation. (More details can be found in Supplementary Video S2, from 0:48 to 1:14 s)

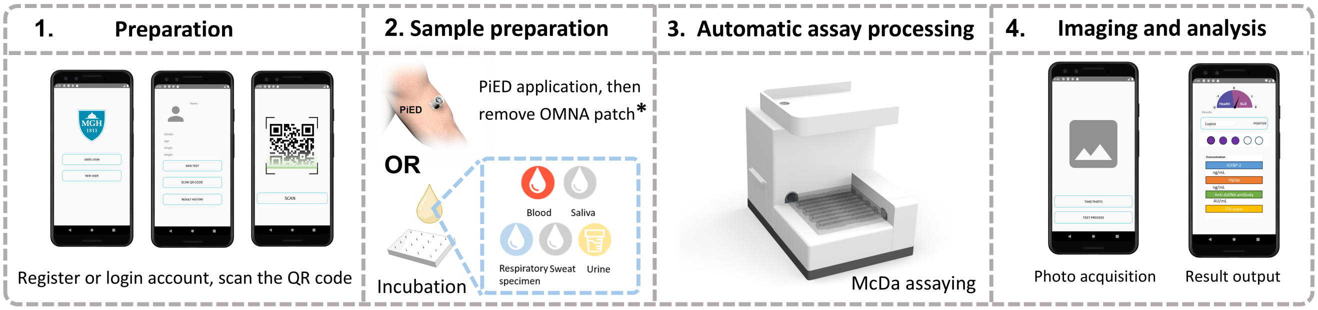


**Supplementary Fig. S15**. Illustration of workflow of miniDia. More detail can be found in Supplementary Video S2.

* More details in *Advanced Materials*, 2025, 37, 27, 2416240**.**

**Supplementary Table S1**. Functionality comparison of commercial ELISA kits with McDa

| Assay | LoD | Sensitivity improvement (v.s. R&D; fold) | Linear Range |
|---|---|---|---|
| Human CRP ELISA Kit (R&D) | 10.14 pg/mL (440.85 fM) | | 15.6-1000 pg/mL |
| **Our work** | **0.47 pg/mL (20.23 fM)** | **21.57** | **0.98-1000 pg/mL** |
| Human IGFBP-2 ELISA kit (R&D) | 12.92 pg/mL (371.16 fM) | | 62.5-4000 pg/mL |
| **Our work** | **0.87 pg/mL (25.21 fM)** | **14.85** | **2.34-1200 pg/mL** |
| Human TNFRII ELISA kit (R&D) | 3.75 pg/mL (189.39 fM) | | 7.81-500 pg/mL |
| **Our work** | **0.21 pg/mL (10.43 fM)** | **17.86** | **0.5-100 pg/mL** |
| Human VCAM-1 ELISA kit (R&D) | 9.15 pg/mL (87.12 fM) | | 15.6-1000 pg/mL |
| **Our work** | **0.30 pg/mL (2.86 fM)** | **30.5** | **0.98-1000 pg/mL** |

**Supplementary Table S2**. Representative results of comparing between McDa and ELISA

| | CRP (µg/mL) | | IGFBP2 (ng/mL) | | TNFRII (ng/mL) | | VCAM-1 (µg/mL) | | Anti-disDNA abs (AU/mL) | |
|---|---|---|---|---|---|---|---|---|---|---|
| | **ELISA** | **McDa** | **ELISA** | **McDa** | **ELISA** | **McDa** | **ELISA** | **McDa** | **ELISA** | **McDa** |
| **Healthy control** | 1.01 | 1.26 | 97.70 | 66.98 | 2.01 | 2.18 | 0.27 | 0.26 | 81.25 | 57.26 |
| **Patient 1** | 13.21 | 14.99 | 218.50 | 397.92 | 12.80 | 19.90 | 0.34 | 0.34 | 217.50 | 259.74 |
| **Patient 2** | 1.19 | 0.50 | 226.62 | 218.07 | 11.07 | 9.63 | 0.52 | 0.48 | 137.00 | 118.54 |
| **Patient 3** | 4.90 | 4.57 | 84.17 | 56.77 | 12.26 | 15.10 | 0.57 | 0.68 | 102.50 | 72.82 |

**Supplementary Table S3.** Spike and recovery of human CRP, IGFBP-2, TNFRII, VCAM-1 in serum samples using McDa

| **Sample** | **Spike level** **CRP** pg/mL | **Expected** pg/mL | **Measured** pg/mL | **Recovery** % |
|---|---|---|---|---|
| Serum | 50 | 48.04 | 47.29±2.6 | 98.43 |
| | 5 | 5.12 | 5.70±0.18 | 111.26 |
| | 0.5 | 0.52 | 0.55±0.03 | 103.77 |

| **Sample** | **Spike level** **IGFBP2** pg/mL | **Expected** pg/mL | **Measured** pg/mL | **Recovery** **%** |
|---|---|---|---|---|
| Serum | 90 | 93.51 | 87.55±2.31 | 93.63 |
| | 9 | 9.36 | 8.48±0.79 | 90.67 |
| | 0.9 | 0.81 | 0.88±0.04 | 104.52 |

| **Sample** | **Spike level** **TNFRII** pg/mL | **Expected** pg/mL | **Measured** pg/mL | **Recovery** % |
|---|---|---|---|---|
| Serum | 20 | 19.73 | 20.46±1.97 | 107.27 |
| | 2 | 2.24 | 2.24±0.14 | 101.97 |
| | 0.2 | 0.19 | 0.18±0.01 | 94.74 |

| **Sample** | **Spike level** **VCAM-1** pg/mL | **Expected** pg/mL | **Measured** pg/mL | **Recovery** % |
|---|---|---|---|---|
| Serum | 30 | 31.41 | 31.36±1.39 | 99.84 |
| | 3 | 2.87 | 3.01±0.19 | 104.76 |
| | 0.3 | 0.33 | 0.29±0.02 | 88.89 |

**Supplementary Table S4. Comparison of multiplex biomarker detection platforms in literature.**

| Summary of function | Sensing type | AI-defined biomarker panel | Clinically improved diagnosis | Home-use potential*, ** and other concerns | Sensitivity | Ref. |
|---|---|---|---|---|---|---|
| Plasmonic protein microarray on gold nano-island film coated slide | Fluorescent | No | Limited<br>*For cardiac troponin I*: Yes; *For creatine kinase isoenzyme MB*: Specificity is lower than that of standard test;<br>No data for biomarker panel | *Low* *<br>*Low convenience***<br>Microarray scanner required | Higher sensitive than ELISA | 1 |
| Protein microarray by inkjet printing of capture antibodies and fluorescently labeled detection antibodies on one chip | Fluorescent | No | N/A | *High**<br>*Low convenience***<br>Safety concern: a 635-nm laser in device | Higher than ELISA | 2 |
| Protein microarray using functionalized single-walled carbon nanotubes (SWNTs) probe | Raman | No | N/A | *Low**<br>*Low convenience***<br>Raman microscope required | Higher sensitive than ELISA | 3 |
| Gold nanorod decorated with bovine serum albumin, biotin, and IRDye 800CW fluorophores (Plasmonic-fluor) for microarray | Fluorescent & colorimetric | No | N/A | *Low**<br>*Low convenience***<br>Microarray scanner required | N/A | 4 |
| Plasmonic-fluor for detection of *single biomarker* on microneedle array | Fluorescent | No | N/A | *Low**<br>*Low convenience***<br>Fluorescence imager | Higher sensitive than ELISA | 5 |
| Protein microarrays using a photoinitiator to form a fluorescent polymeric film by light induced polymerization | Colorimetric & fluorescent | No | N/A | *Moderate to Low**<br>*Low convenience***<br>*For profilometry:* Surface profilometer<br>*For fluorescent:* Microarray scanner | *For profilometry or Digital camera imaging*: less sensitive than ELISA<br>*For Microarray scanner:* comparable to ELISA | 6 |
| Protein microarray using antibody functionalized gold nanorods | Colorimetric | No | N/A | *Low**<br>*Low convenience*** | Higher sensitive than ELISA | 7 |

| | | | | | | |
|---|---|---|---|---|---|---|
| | | | | Microscope required | | |
| Protein microarray using antibody and oligonucleotide modified Au nanoparticles (NPs) and Au NPs initiated gold deposition | Sconometric | No | N/A | *Low**<br>*Low convenience***<br>Light scattering reader system | Higher sensitive than ELISA | 8 |
| Protein microarray using HRP initiated silver amplification | Sconometric | No | N/A | *Moderate**<br>*Low convenience***<br>Flatbed scanner | Some are comparable to ELISA<br>Some are less sensitive than ELISA | 9 |
| Gold electrode coated by bovine serum albumin and reduced graphene oxide nanoflakes (rGOx) crosslinked with glutaraldehyde | Electrochemical | No | N/A | *Moderate**<br>*Low convenience***<br>Potentiostat required | Less sensitive than ELISA | 10 |
| Gold nanostructured microelectrodes tethered aptamer/ferrocene probe | Electrochemical | No | N/A | *Moderate**<br>*Low convenience***<br>Potentiostat required | Higher sensitive than ELISA | 11 |
| Mobile digital droplet ELISA platform using parallelized microfluidic droplet generators and cellphone-based fluorescence imaging | Fluorescent | No | N/A | *High**<br>*Low convenience***<br>Limitations: Limited number of biomarker detection due to fluorescent dye labels; lasers with different excitation lights needed;<br>Safety concern: Lasers in device | Higher sensitive than ELISA | 12 |
| lateral flow assay (LFA) based on silver core and gold shell NPs surface enhanced Raman scattering (SERS) nanotags | Raman | No | N/A | *Low**<br>Raman microscope required | Higher sensitive than ELISA | 13 |
| **iPlexD2Go** | **Colorimetric** | **Yes** | **Yes** | **Smart phone for imaging miniDia (cost: < US$180) Automatic device** | **Higher than ELISA** | **This work** |

*Home-use potential was categorized into three criteria mainly according to the instrument/device cost required for testing (because this cost is critical for home-use potential): (1) **High**, cost < US$300 (e.g., miniDia); (2) **Moderate**, cost US$1,000-10,000 (e.g., surface profilometer, flatbed scanner, and potentiostat); (3). **Low:** cost > US$10,000 (e.g., microarray scanner, Raman microscope, fluorescence imager, optical microscope, and light scattering reader system).

** Low convenience: No automatic operation is available for sample and reagent loading, and skilled personnel are needed.

**Supplementary Video S1.** The illustration of 3D-to-2D dimensional conversion for signal amplification.

**Supplementary Video S2. The workflow, architecture, components, and working principles of miniDia and McDa.** The video shows that the process begins with scanning a QR code to register the test, followed by connecting the reagent-prefilled millifluidic cartridge and inserting the MNA patch and the substrate-saturated gel (0–24 s). Second, the miniDia device automatically performs sequential washing and assaying using an MCU-controlled, on-demand vacuum system. Driven by a micro-stepper actuator, the vacuum sequentially draws preloaded reagents from the millifluidic cartridge into the immunoassay chamber without manual intervention (0:48–1:14). Third, it illustrates the detailed operating mechanism of the McDa assay (1:15–2:17) and signal conversion, imaging, and data analysis (2:17–3:03). After the immunoassaying, a micro-stepper motor drives the MNA into the substrate-saturated imaging gel, converting the 3D MNA signals into a 2D color pattern for smartphone imaging under standardized LED illumination. The acquired images are then analyzed using a deep-learning algorithm, and the final diagnostic output is generated by a machine-learning model.
